# Cancer driver genes and opportunities for precision oncology revealed by whole genome sequencing 10,478 cancers

**DOI:** 10.1101/2023.05.24.23289454

**Authors:** Ben Kinnersley, Amit Sud, Andrew Everall, Alex J. Cornish, Daniel Chubb, Richard Culliford, Andreas Gruber, Adrian Lärkeryd, Costas Mitsopoulos, Genomics England Research Consortium, David Wedge, Richard Houlston

## Abstract

Identifying cancer driver genes is key for delivering the vision of precision oncology. The falling cost of whole genome sequencing (WGS) potentially makes WGS an attractive single all-encompassing test to identify cancer drivers in a patient, which may not be captured by standard panel testing but are targetable by small molecules. We analysed WGS data on 10,478 patients spanning 35 cancer types recruited to the UK 100,000 Genomes Project. We identified 330 driver genes, including 74 which are novel to any cancer. Across all cancer types 16% of the patients would be eligible for a currently approved therapy. Computational chemogenomic analysis of cancer mutations identified 96 additional targets of compounds that are potentially active and represent candidates for future clinical trials, expanding opportunities for improved patient care.

## INTRODUCTION

The rationale for the one-size-fits-all medical treatment model is being challenged with the move towards individualised therapy^1^. This is epitomised in oncology where standard therapies are reported to be ineffective in around 75% of patients, representing one of the highest therapy failure rates in all diseases^2,3^.

Precision oncology describes a set of strategies tailored to the unique biology of a patient’s disease. The potential promise of this approach includes improved treatment efficacy, more favourable toxicity profiles and a reduction in the administration of ineffective treatments^4^. Underpinning precision oncology is the concept of somatic driver mutations as the foundation of cancer development^5^. There are already a number of approved therapies for tumours with specific “actionable” driver mutations, with additional ones in development^6^. Knowledge of the actionable driver mutational landscape in cancers has recently become central to delivering precision oncology.

Currently multiple standalone tests or a panel are typically used to capture a set of genomic features for a given tumour type. However, falling costs make whole genome sequencing (WGS) a potentially attractive proposition as a single all-encompassing test^7^. Moreover, it provides the opportunity to identify additional cancer drivers in a patient which may not be captured by standard clinical panel tests. This affords the opportunity to broaden the scope of cancers potentially amenable to small molecule therapies. To examine this proposition in the real-world setting we analysed WGS data from 10,478 cancer patients recruited to the Genomics England (GEL) 100,000 Genomes Project (100kGP) (**Fig 1A**)^8^.

**Figure 1.**
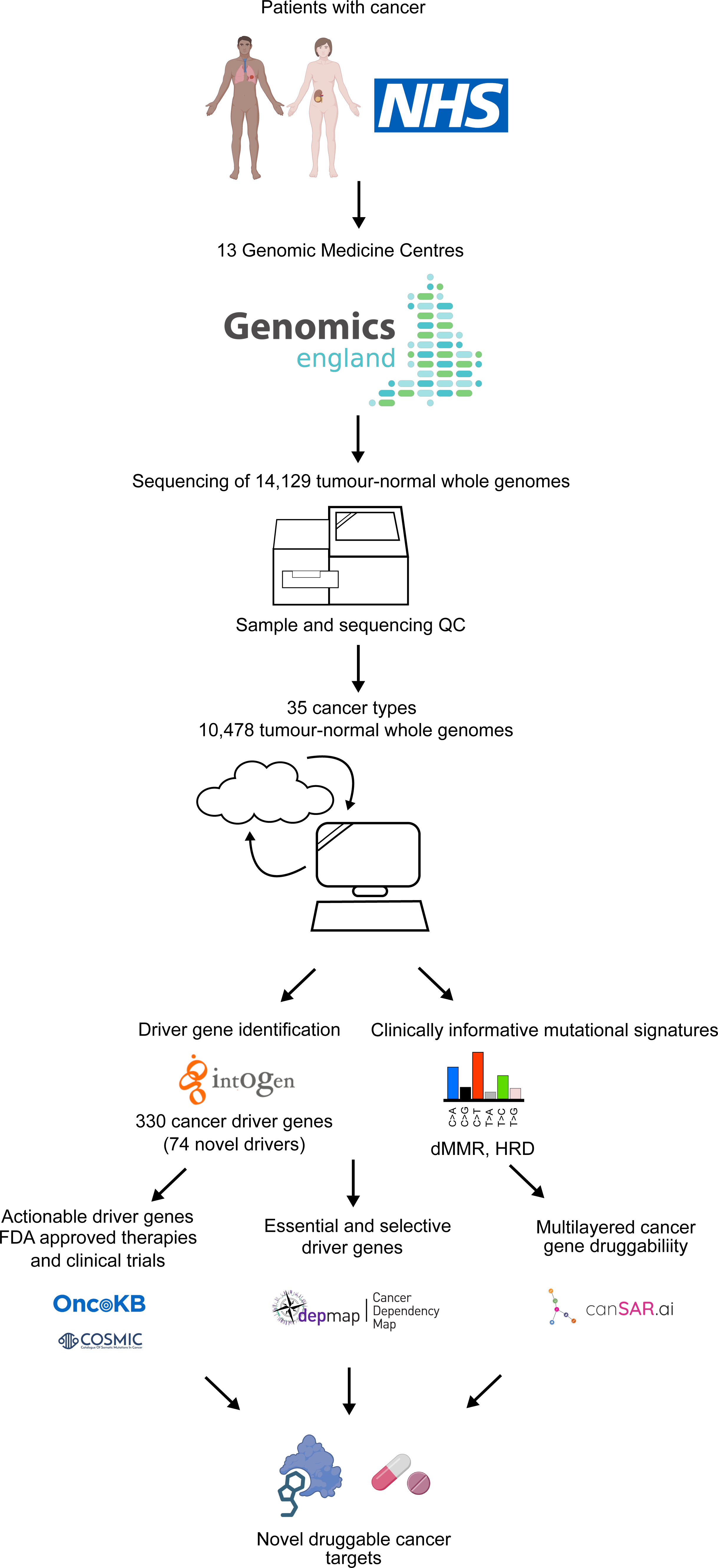
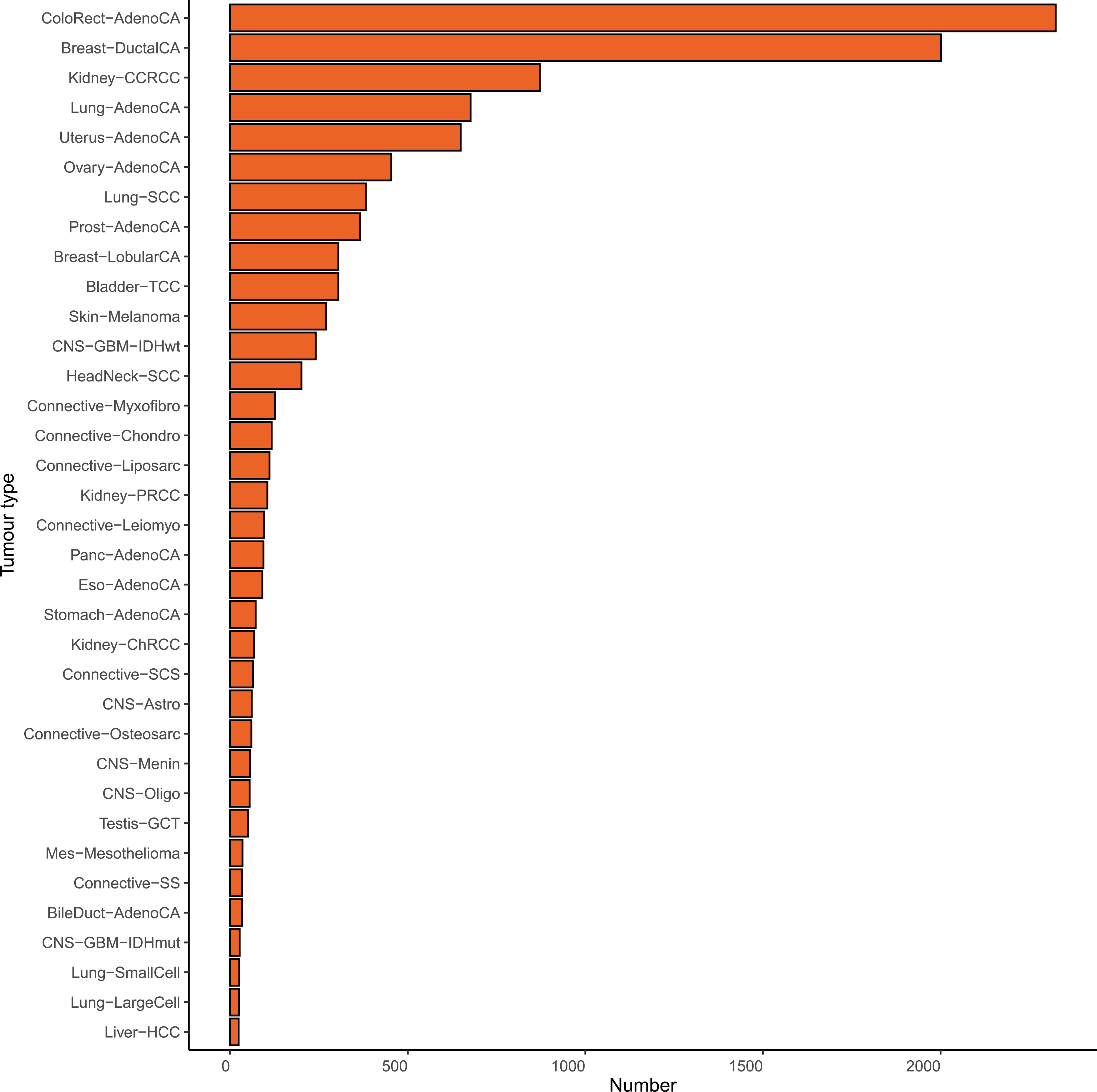
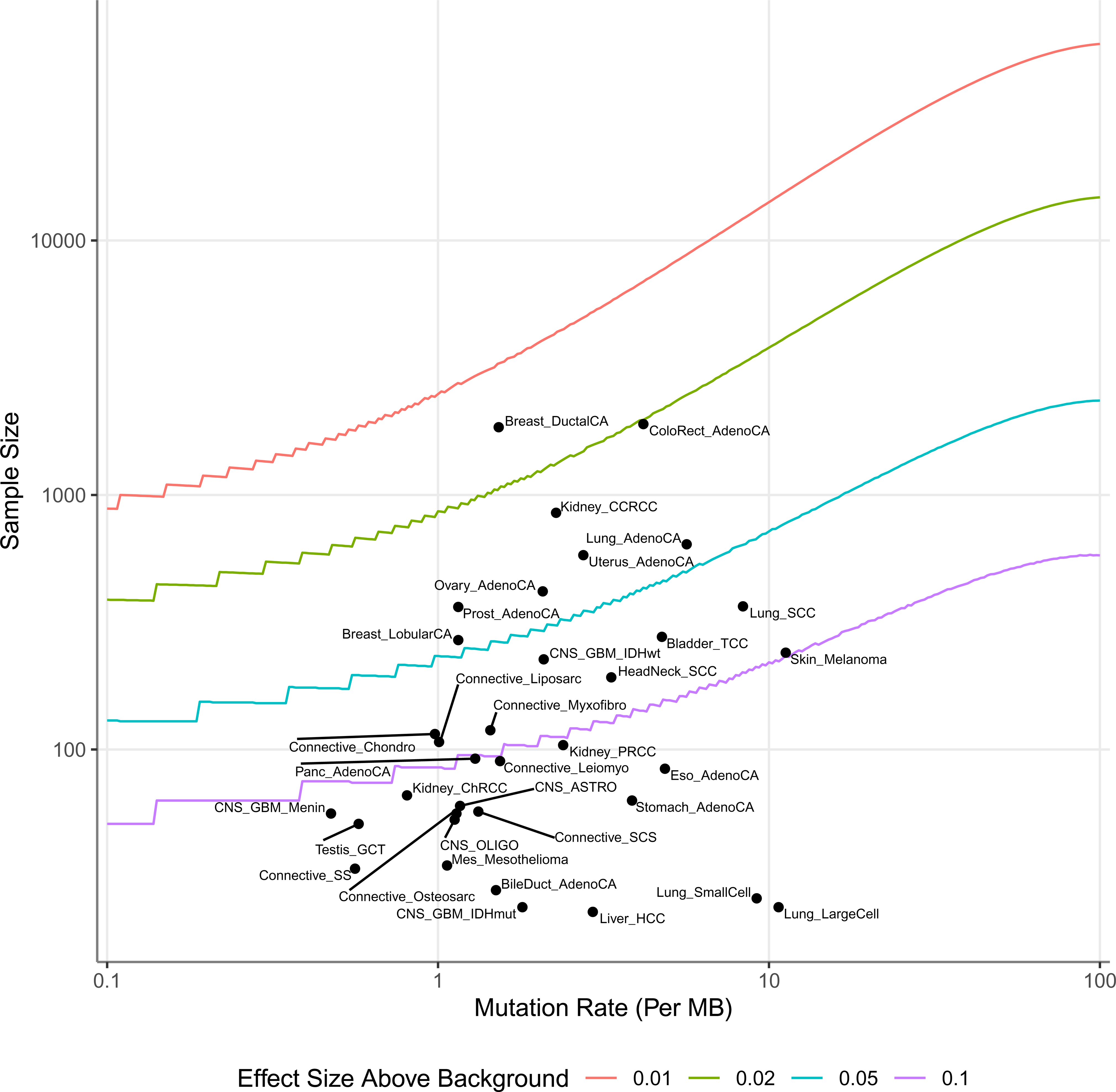
(A) Study design; (B) Number of samples per tumour type; (C) Power estimates for driver gene identification per tumour type. The number of samples needed to achieve 90% power for 90% of genes (*y* axis). Grey vertical lines indicate exome-wide background mutation rates (*x* axis). Black dots indicate sample sizes in the current study. For most tumour types, the current sample size is inadequate to reliably detect genes mutated at 5% or less above background. BileDuct-AdenoCA, cholangiocarcinoma; Bladder-TCC, bladder transitional cell carcinoma; Breast-DuctalCA, breast ductal carcinoma; Breast-LobularCA, breast lobular carcinoma; CNS-Astro, astrocytoma; CNS-GBM-IDHmut, IDH mutated glioblastoma; CNS-GBM-IDHwt, IDH wild type glioblastoma; CNS-Menin, meningioma; CNS-Oligo, oligodendroglioma; ColoRect-AdenoCA, colorectal adenocarcinoma; Connective-Chondro, chondrosarcoma; Connective-Leiomyo, leiomyosarcoma; Connective-Liposarc, liposarcoma; Connective-Myxofibro, myxofibrosarcoma; Connective-Osteosarc, osteosarcoma; Connective-SCS spindle cell sarcoma; Connective-SS, synovial sarcoma; Eso-AdenoCA, esophageal adenocarcinoma; HeadNeck-SCC, squamous cell carcinoma of the head and neck; Kidney-CCRCC, clear cell renal cell carcinoma; Kidney-ChRCC, chromophobe renal cell carcinoma; Kidney-PRCC, papillary renal cell carcinoma; Liver-HCC, hepatocellular carcinoma;Lung-AdenoCA, lung adenocarcinoma; Lung-LargeCell, large-cell lung cancer; Lung-SCC, squamous cell carcinoma of the lung; Lung-SmallCell, small cell carcinoma of the lung; Mes-Mesothelioma, mesothelioma; Ovary-AdenoCA, ovarian adenocarcinoma; Panc-AdenoCA, pancreatic adenocarcinoma; Prost-AdenoCA, prostate adenocarcinom; Skin-Melanoma, melanoma of the skin; Stomach-AdenoCA, gastric adenocarcinoma; Testis-GCT, testicular germ cell tumour; Uterus-AdenoCA, uterine adenocarcinoma.

## METHODS

### The GEL cohort

We restricted our analysis to high-quality data derived from PCR-free, flash-frozen primary solid tumour samples from adults (v8 data release), resulting in 10,478 samples (34 bile duct, 305 bladder, 2,306 breast, 2,324 colorectal, 440 central nervous system, 91 esophageal, 201 head and neck, 1,045 renal cell, 24 liver, 1,466 lung, 35 mesothelioma, 607 soft-tissue, 454 ovarian, 94 pancreas, 366 prostate, 270 melanoma, 72 gastric, 51 testicular, 649 uterus) from 10,470 individuals (**Supplementary Tables 1-3**). Complete details on sample curation, WGS, somatic variant calling, mutation annotation and power calculations are provided in **Supplementary Methods**.

### Identification and actionability of driver genes

Cancer driver genes were identified using IntOGen^9^, which combines seven computational methods to detect signals of positive selection from the mutational patterns of genes in each cancer type (**Supplementary Methods**). Details of pre-processing of mutations, combining driver gene identification methodologies, post-processing and annotation of driver gene mutations are provided in **Supplementary Methods**. Information on the clinical actionability of cancer drivers were retrieved by querying OncoKB^6^ and COSMIC^10^.

### Sensitivity of WGS

We tested the sensitivity of WGS in 100kGP to detect driver gene mutations based on sample purity and gene coverage and by comparing call rates of panel sequencing reported in the IMPACT and MET studies of cancer conducted by Memorial Sloan Kettering Cancer Center (MSK) (**Supplementary Methods)**^11,12^.

### Mutation signature analysis

Tumours with microsatellite instability (MSI) were identified using MSINGS^13^ and homologous recombination deficiency was assessed using HRDetect^14^. Further details are provided in the **Supplementary Methods.**

### Chemogenomic annotation of cancer networks

To construct networks for each cancer type, we used protein products of the cancer driver genes to seed a search for all interacting proteins in the canSAR interactome^15^, which is based on information from 8 databases, including the IMeX consortium^16^, Phosphosite^17^, and key publications. The canSAR interactome features interactions where there are: (i) ≥2 publications with experimental evidence of binary interaction between the two proteins; (ii) 3D protein evidence of a complex; (iii) ≥2 reports that one protein is a substrate of the other; (iv) ≥2 publications reporting that one protein is the product of a gene under the direct regulatory control of the other. Each tumour-specific interactome was seeded using cancer driver proteins retrieving interacting proteins that had supporting experimental evidence. To ensure only additional proteins are likely to function primarily through interaction with proteins in the network we adopted the following strategy: Starting with the input list of proteins we obtained all possible first neighbours. We then computed, for each new protein, the proportion of its first neighbours in the original input list. To define proteins likely to function through the network, we calculated the probability of these occurring randomly, by permuting the interactome 10,000 times. We corrected empiric *P*-values for multiple testing retaining only proteins having a FDR < 0.05. For each cancer type we minimised the network by retaining only proteins connected to more than one cancer protein, or whose only connection was to a cancer-specific protein. We then annotated proteins with pharmacological and druggability data using canSAR’s Cancer Protein Annotation Tool (CPAT). Essential and selective genes including lineage specificity were ascertained from the ShinyDepMap analysis^18^.

## RESULTS

We analysed genomes from 10,478 cancers comprising 35 different cancer types (**Fig. 1B** and **Supplementary Table 2**). While broadly reflecting the spectrum and frequencies of cancers diagnosed in the UK population there were differences, with an over-representation of colorectal and kidney cancers and paucity of prostate and pancreatic cancers (**Supplementary Fig. 1**). It was also the case that for the major cancer types, patients recruited to 100KGP tended to be younger and had earlier stage cancers than the general population (**Supplementary Table 3**).

Mutation rates varied across the different cancer types with cutaneous melanoma having highest single nucleotide variant (SNV) mutation count and meningioma the lowest SNV mutation count (**Supplementary Fig. 2)**. 945 samples, notably colorectal and uterine cancers, were hypermutated, either as result of deficient mismatch repair (dMMR) or *POLE* mutation. Invasive ductal carcinoma of the breast had the highest power for driver gene detection (>90% power for a mutation rate of at least 2% higher than background) and LCLC the lowest power (**Fig. 1C** and **Supplementary Table 4**). Compared with the recent PCAWG pan-cancer analysis^19^, the 100kGP cohort was better powered to identify a driver mutation for 19 cancers. Notably, in breast, colorectal, oesophageal and uterine cancer, lung adenocarcinoma and bladder transitional cell carcinoma where the sample sizes were >10-fold higher.

### Spectrum of cancer driver genes

Across all cancer types we identified 770 unique tumour-driver gene pairs corresponding to 330 unique cancer driver genes (**Fig. 2A** and **Supplementary Table 5**). When compared to the largest pan-cancer driver analysis, in 21 of 31 cancer types with matching tumour histologies we recovered 61% of all cancer drivers reported by COSMIC, IntOGen and the TCGA pan-cancer analysis reported in Bailey *et a l*,. 2018 (**Supplementary Table 5**)^9,20^. We were able to detect 80% of drivers reported for colorectal, breast, lung and ovarian cancers but only <20% of drivers for hepatocellular and stomach cancers, which is likely a result of differing sample size. The number of identified cancer driver genes varied between cancer types, with colorectal and uterine cancers having the most (60 genes) and spindle cell carcinoma having the fewest (4 genes). Across the 35 cancers, we found no correlation between average mutation burden and the number of driver genes in each cancer (Pearson R=0.19, *P*-value=0.27). The consensus list also includes 330 tumour-driver pairs that have not previously been reported by either the Cancer Gene Census (CGC), Intogen or the PanCancerSoftware analysis of TCGA^9,20^ (**Supplementary Table 5**), and 74 that have not previously been associated with any specific tissue. Almost all of the novel drivers identified were uncommon with 65/74 (88%) possessing a mutated frequency <10% in each cancer type. We observed the highest number of new cancer driver genes for uterine (n=42), bladder (n=40) and colorectal (n=37) cancers. Moreover, we are able to identify drivers in tumour types which have been relatively unexplored by the Intogen and PanCancerSoftware catalogues^9,20^. These include breast lobular carcinoma, meningioma and myxofibrosarcoma. Predictions of known cancer driver genes in new cancer types include *SPTA1*, *CHD4* and *ASXL1* in colorectal cancer, *FOXO3*, *MUC16* and *ZFPM1* in breast cancers and *CNTNAP2*, *CTNND2* and *TRRAP* in lung adenocarcinoma. Entirely novel predictions include *MAP3K21* (mixed-lineage kinase) in colorectal cancer, *USP17L22* (deubiquitinating enzyme) in breast ductal carcinoma, and *TPTE* (tyrosine phosphatase) in lung adenocarcinoma (**Supplementary Table 5**).

**Figure 2.**
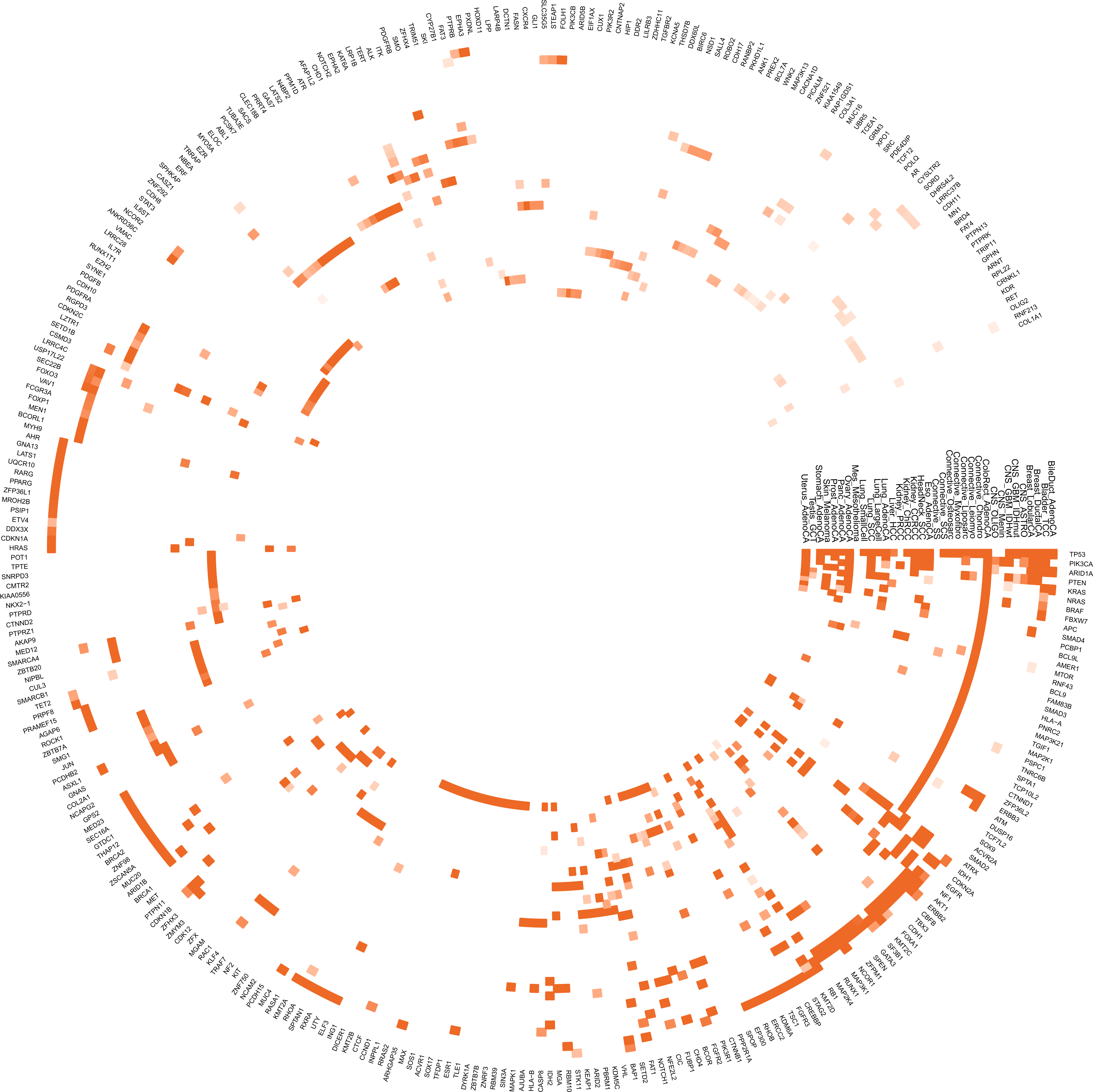
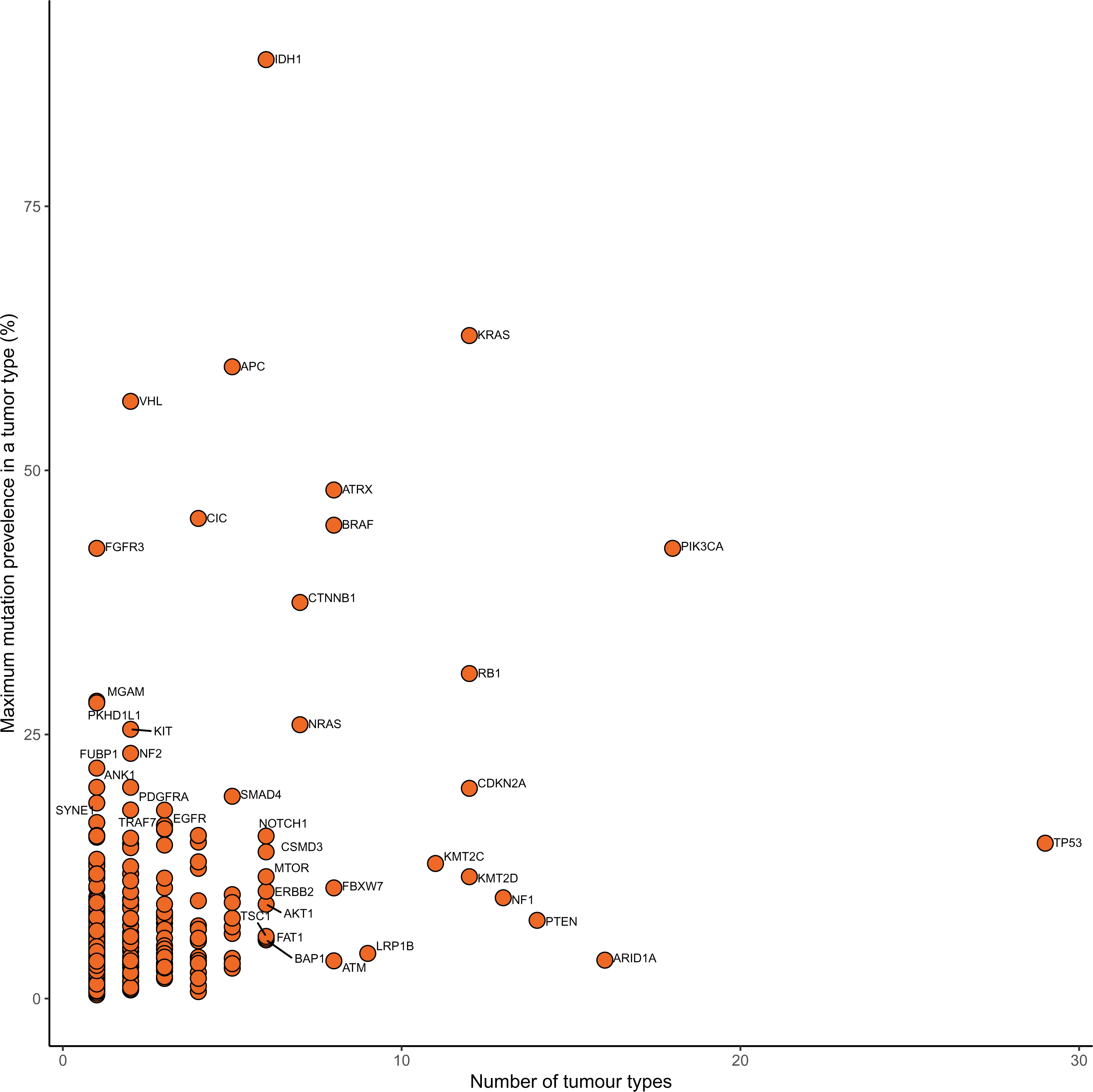
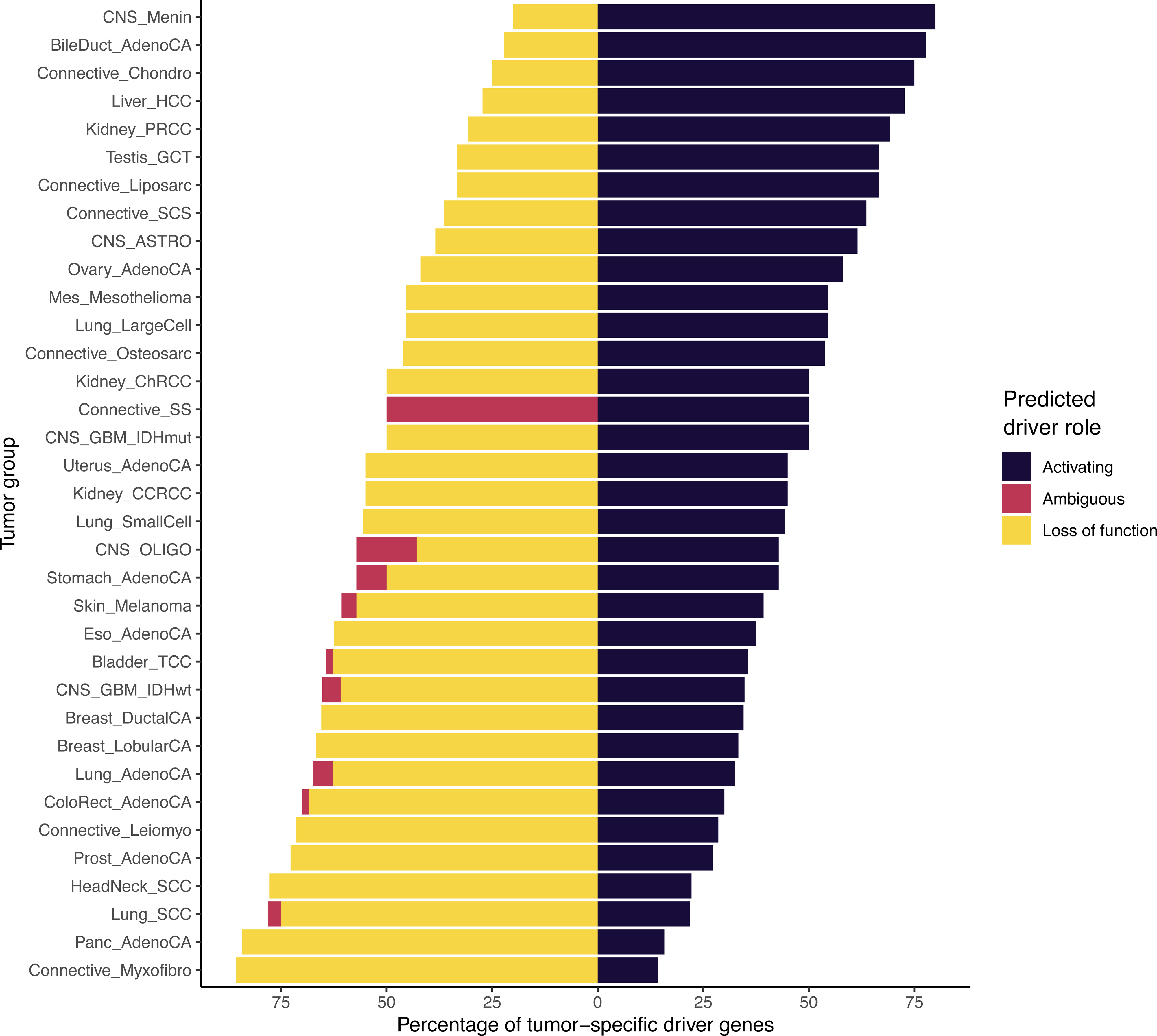
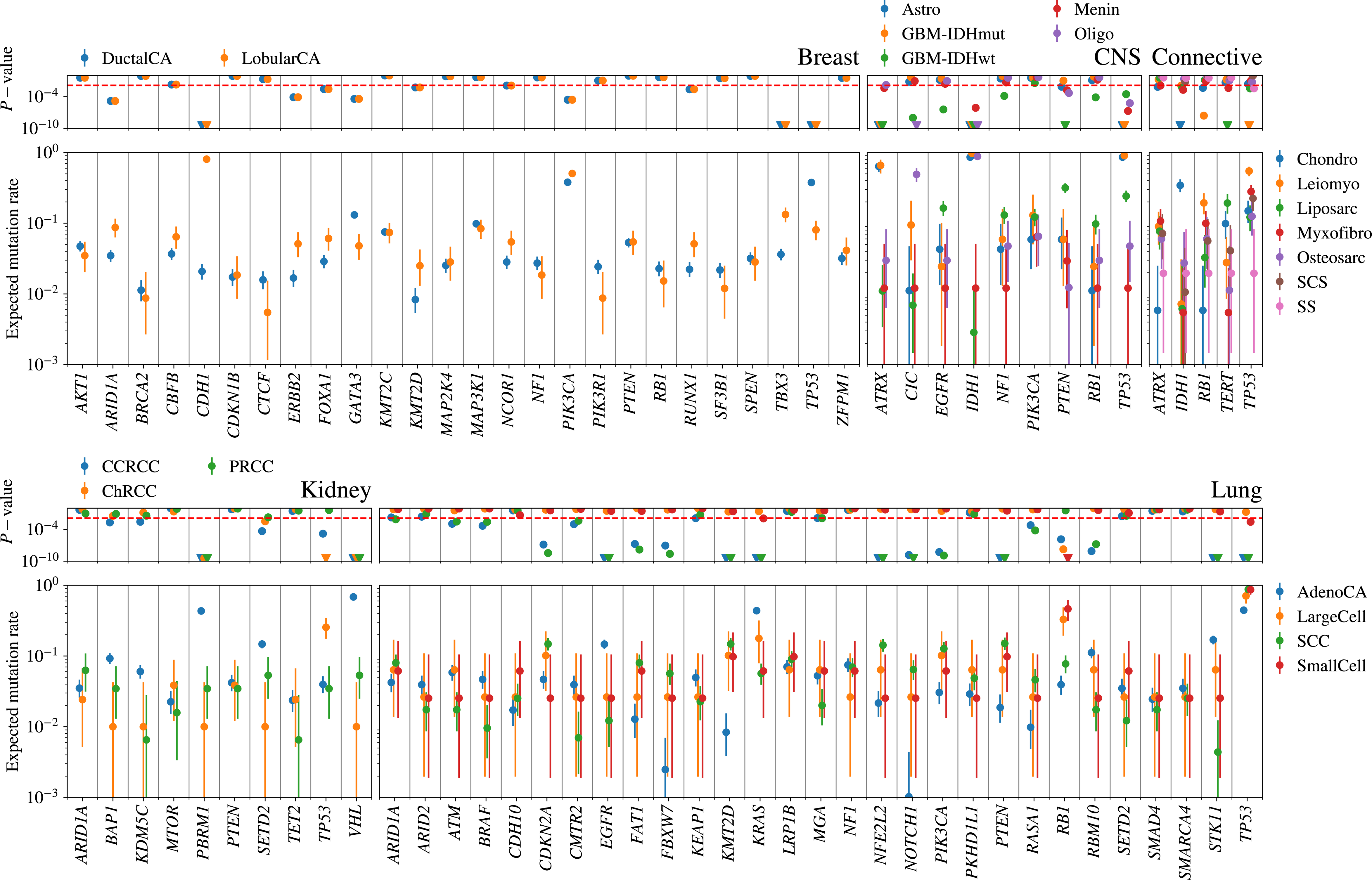
(A) Circos heatmap of cancer driver genes identified. Heatmap intensity proportional to Stouffer *P*-value. **(B) Distribution of driver genes across different types of cancer.** y-axis, maximal mutational prevalence in a tumour type, x-axis, number of tumour types in which the driver gene is identified. **(C) Distribution of cancer driver gene function associated with each cancer type.** Y-axis, tumour group, x-axis, percentage of tumour specific driver genes. **(D) Comparison of driver gene somatic mutation rates between tumour histologies.** Expected mutation rate of each driver in the cohort based on the number of samples in which the driver gene is mutated for the given tumour histology. Binomial *P*-values are shown.

Considering the prevalence of driver genes across cancer types, some genes were seen to act as drivers across multiple cancer types, while others more specific. Eighty-five genes were identified as a driver in more than two tumour types, with 26 genes functioning as drivers in more than five tumour types (**Fig. 2B**). As expected, *TP53* was identified as a driver gene in the largest number of tumour types, followed by *PIK3CA*, *ARID1A* and *PTEN*, acting as cancer driver genes in 29, 18, 16 and 14 different tumour types respectively. While many genes function as drivers in multiple cancer types, some drivers are mutated at high frequencies only in specific tumours, such as *VHL* in clear cell renal cell carcinoma and *FGFR3* in bladder cancer (**Fig. 2B**). Across drivers operating in multiple cancer types, the clearest example of domain specific driver mutations were in *EGFR,* where protein tyrosine and serine/threonine kinase domain mutations predominated in lung adenocarcinoma, in contrast to extracellular furin-like cysteine rich region domain mutations in IDH wild-type glioblastoma (**Supplementary Table 6** and **Supplementary Fig. 3a-b)**. At PIK3CA we also observed a preference for p85 binding domain mutations in uterus adenocarcinoma compared to other cancer types, such as breast ductal carcinoma, which are enriched for mutations in the PIK family domain (**Supplementary Table 6** and **Supplementary Fig. 3c-d**). Hierarchical clustering of cancers based on the presence of identified driver mutations and their respective *Q*-value demonstrated clustering of cancer types by cell of origin (*e.g.* head and neck and lung squamous cell carcinoma) and by organ (*e.g.* breast ductal and lobular carcinomas) (**Supplementary Fig. 4**). The ratio of predicted activating to tumour suppressor driver genes varied across tumour types with meningioma and myxofibrosarcoma possessing the highest and lowest ratio respectively (**Fig. 2C** and **Supplementary Table 5**).

Across the 35 different tumour types we identified 12,606 distinct oncogenic mutations in tumour-relevant cancer driver genes, in 9,070 unique samples. The median number of oncogenic mutations in cancer driver genes per sample was 2 across all tumours. The highest median number of oncogenic mutations in cancer driver genes per sample was seen in uterine cancer (**Supplementary Fig. 5**). We observed significant differences in oncogenic mutation frequency in cancer driver genes across different tumour histologies arising from the same organ. Examples include *CDH1*, *TBX3* and *TP53* in breast cancers, *ATRX*, *CIC*, *IDH1*, *PTEN* and *TP53* in central nervous system tumours, *IDH1* and *TP53* in connective tissue tumours, *PBRM1* and *VHL* in renal cancers and *EGFR*, *KMT2D*, *KRAS*, *NFE2L2*, *PTEN*, *STK11* and *TP53* in lung cancer (**Fig. 2D)**.

Of the 330 cancer driver genes, 214 possessed at least one oncogenic mutation annotated as clonal, 167 as early and 114 as late events (**Supplementary Table 7**). APC, *TP53* and *PIK3CA* possessed the highest number of clonal oncogenic mutations. Of the 162 driver genes that harboured at least one subclonal oncogenic mutation, *ARID1A*, *TP53* and *PIK3CA* possessed the highest number (**Supplementary Fig. 6**). In tumours with >10 oncogenic mutations, meningioma possessed the greatest proportion of clonal oncogenic mutations (**Supplementary Fig. 7a**). Large-cell lung, testicular germ cell tumour and oligodendroglioma carried the highest proportion of early clonal, late clonal and subclonal oncogenic mutations respectively (**Supplementary Fig. 7b-d**).

### Sensitivity of WGS mutation detection as compared to cancer gene panels

For primary tumours represented in the MSK and 100kGP cohorts, the rate of mutations called for each driver gene were comparable (**Supplementary Fig. 8 and 9**). In 88% of cancer driver genes, the expected sensitivity for mutation detection was >99% in the 100kGP cohort. In 90% of cancer driver genes >98% of the coding sequence had sufficient coverage such that >6 reads could be used for mutation detection (**Supplementary Fig. 10-14**). These findings are in agreement with internal testing of panel sequencing compared to WGS at Genomics England (sensitivity of 99% for VAF > 5% and coverage >70x).

### Clinical implications of genomic features

Systematic analyses of cancer genomes provide an opportunity of estimating the number of individuals eligible for a targeted therapy and identify potentially novel therapeutic interventions. We first used two different databases to evaluate the therapeutic implications of the genetic events: Precision Oncology Knowledge Base (OncoKB) and the COSMIC Mutation Actionability in Precision Oncology Product^6,10^. Both databases catalogue approved marketed drugs having demonstrated efficacy in tumours with specified driver gene mutations, based on clinical trials and published clinical evidence. OncoKB also provides compelling biological evidence supporting the cancer driver gene as being predictive of a response to a given drug.

We observed that both the fraction of samples and proportion of alteration types varied across tissue types. Data from COSMIC indicates that 85% of all samples (8,874/10,478) possess at least one putatively actionable alteration being targeted in a clinical setting (**Fig. 3A** and **Supplementary Table 8**), while 55% of samples (5,805/10,478) had at least one putatively actionable or biologically relevant alteration from OncoKB (**Fig. 3B** and **Supplementary Tables 9** and **10**). Across all cancer types, 16% (1,633/10,470) of the patients would be eligible for a currently approved therapy as defined by OncoKB. Of the actionable mutations defined by OncoKB (n=9,639), 5,823 were clonal, 2,632 were early clonal, 229 were late clonal and 852 were subclonal.

**FIgure 3.**
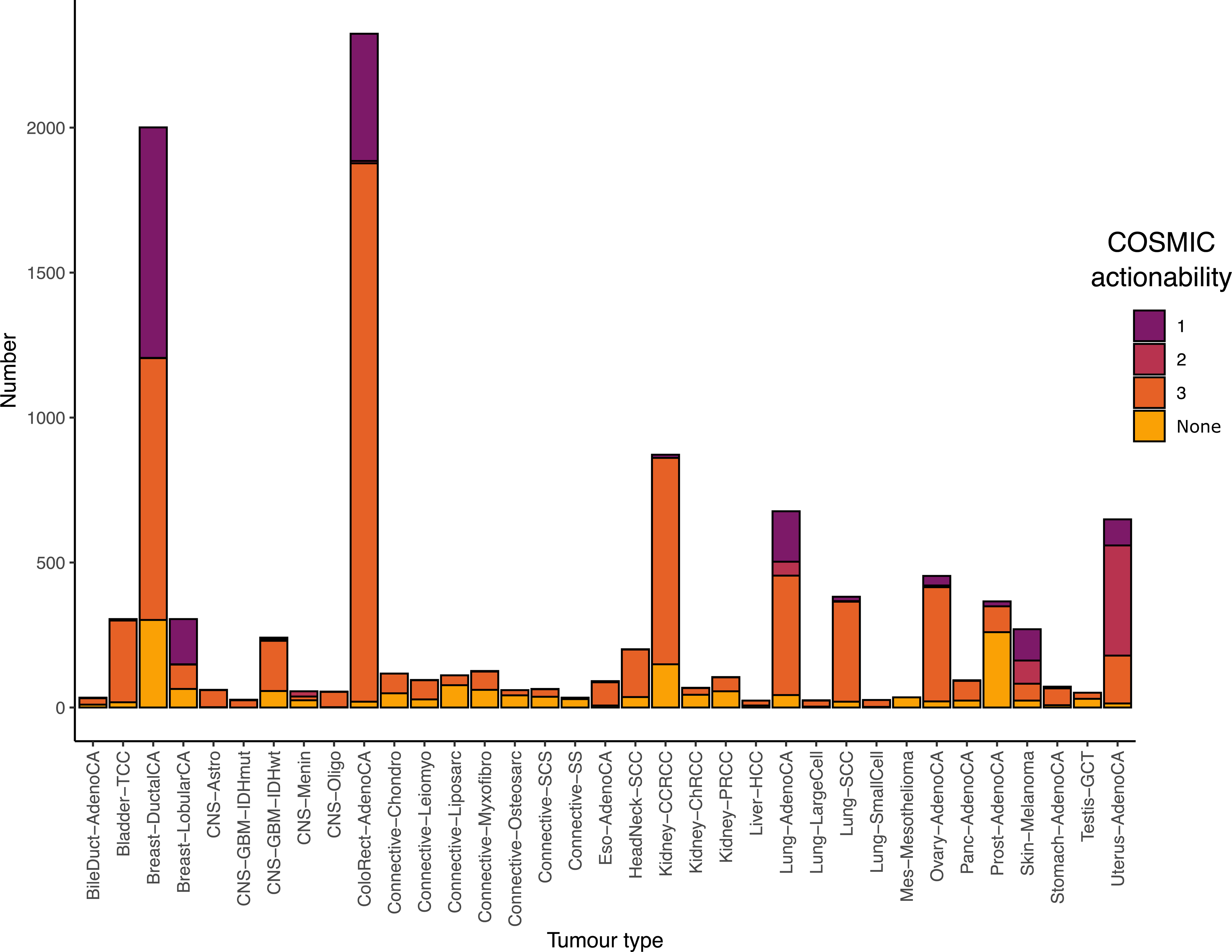
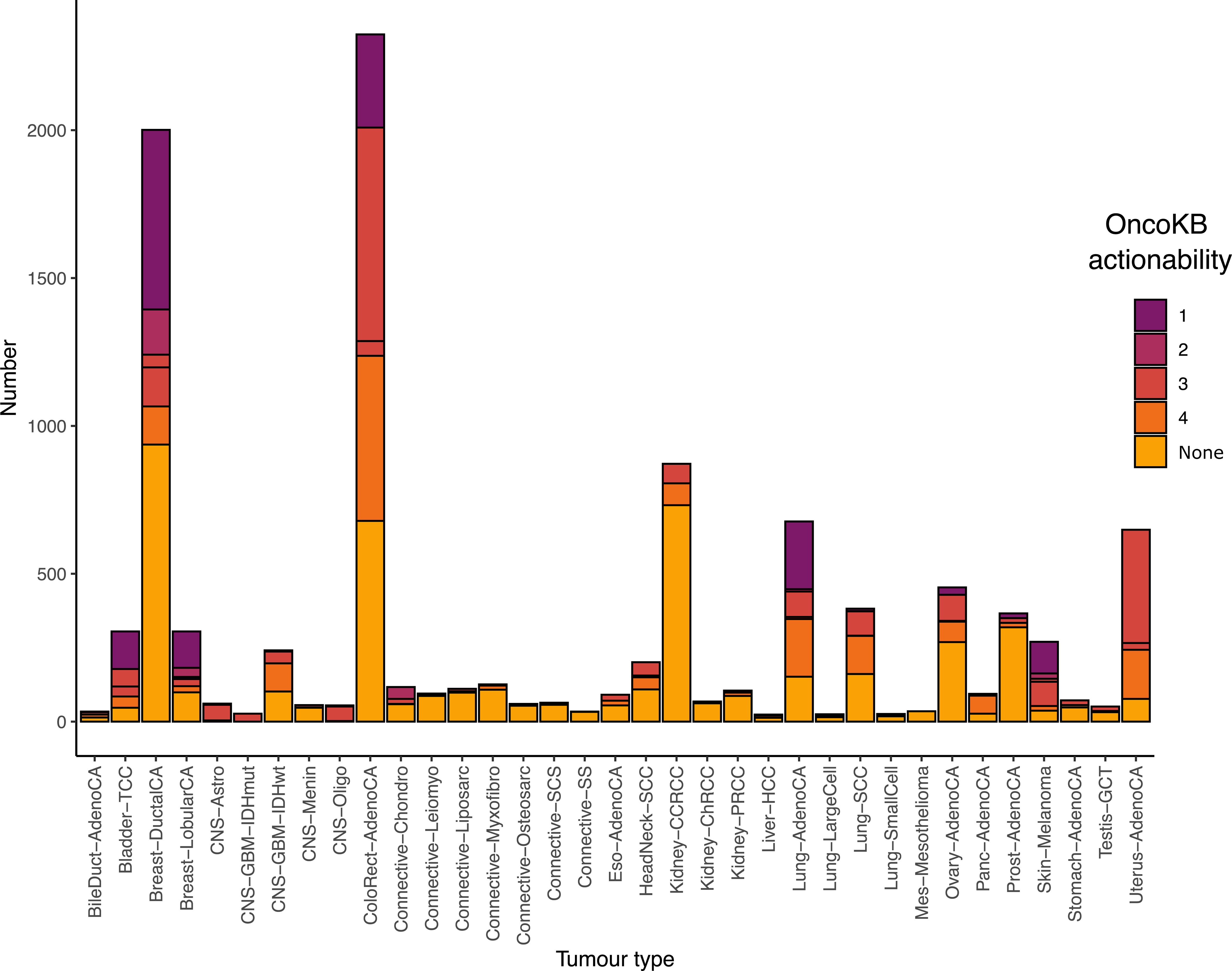
Clinical actionability ascribable to each cancer driver gene according to COSMIC and OncoKB by cancer type. Tumours were annotated by the highest scoring gene mutation - indication pairing, with “None” indicating no actionable mutations were detected in the tumour. **(A) Catalogued by COSMIC:** 1, Approved marketed drug with demonstrated efficacy at the mutation; 2, Phase 2/3 clinical results meet primary outcome measures; 3 Drug in ongoing clinical trials. **(B) Catalogued by OncoKB:** 1, FDA approved drug in the cancer type; 2, standard of care in the cancer type; 3, clinical evidence in the cancer type or standard of care in a different cancer type; 4, supported by compelling biological evidence.

The most common putatively actionable alterations across all of the 35 cancer types were mutations in *PIK3CA*, *KRAS* and *PTEN*. Specific oncogenic missense mutations in *PIK3CA* are present in 40% of lobular breast cancers and 30% of ductal breast cancers and their presence are an indication for the use of PI3Kα-inhibitor Alpelisib^21^. These mutations are present in a number of cancers including colorectal (20%) and uterine cancers (47%) and are subject to early clinical studies with an allosteric inhibitor of PI3Kα. We found high fractions of patients with pancreatic, colorectal cancers and lung adenocarcinoma with actionable *KRAS* mutations (between 34% and 69% of all cases). The G12C mutation was present in 17% of lung adenocarcinoma cases and is targeted by mutation specific selective covalent inhibition with Adagrasib or Sotorasib^22,23^. PI3Kβ inhibition is of significant biological interest in patients with oncogenic *PTEN* mutations as PI3Kβ is thought to drive cellular proliferation in these tumours. These PTEN mutations were prevalent in melanoma (10%), hepatocellular carcinoma (13%), squamous cell carcinoma of the lung (15%), glioblastoma multiforme (29%) and uterine carcinoma (66%) and their presence would result in eligibility for early studies of PI3Kβ inhibition^24^.

319 tumours possessed a HRD mutational signature, providing support for the use of a PARP inhibitor in these individuals. Furthermore, 1,309 tumours possessed a high coding tumour mutational burden (≥10 mutations/megabase) and 144 cancers had evidence of dMMR^6,25^. Considering these collectively would suggest that 1,312 patients may be eligible for checkpoint inhibition. To explore the prospect of multiple targeted therapies being used in the same patient, we combined the OncoKB clinical actionability annotations with that of TMB, dMMR and HRD clinical actionability annotations. In total, 11,534 independent gene targets were present with 28% (2,941/10,478) possessing one, 8% two (828/10,478), and 6% (594/10,478) possessing >3 clinically actionable driver mutations.

Controlling for age, sex and tumour, oncogenic *TP53* mutations were associated with worse survival in breast ductal carcinoma and (*P*=6.4 x 10^-6^, HR=2.59 (95% CI: 1.71-3.92) and oncogenic *KRAS* mutations in colorectal cancer (*P*=1.1 x 10^-3^, HR=1.43 (95%CI: 1.17,1.77) (**Supplementary Fig. 15**).

### Expanding the druggable cancer genome

An opportunity emerging from the systematic analysis of cancer genomes is the identification of novel therapeutic intervention strategies. Of the 330 cancer driver genes identified in this study, 261/330 (79%) are not currently identified as targets in either COSMIC or OncoKB databases. As a means of triaging these genes as candidates for therapeutic intervention, we assessed the essentiality and selectivity of driver genes and their druggability using RNAi/CRISPR DepMap data and the integrative cancer-focused knowledgebase, canSAR respectively. We found 96/261 (37%) of these genes are predicted to be commonly essential and of these 12/96 (13%) have a chemical probe available and 35/96 (36%) have a ligandable 3-dimensional structure (**Supplementary Table 11**).

Motivated by the observation that targeting proteins that interact with cancer driver genes can result in successful precision oncology strategies, we sought to expand the network of druggable targets in cancer. To this end, we used canSAR to map and pharmacologically annotate networks of the cancer genes identified for each tumour type. Specifically, we seeded networks with driver genes identified in each tumour group and used transcriptional and curated protein-protein interactions to recover a refined cancer specific-network of proteins, each protein being annotated based on multiple assessments of ‘druggability’, *i.e*. the likelihood of the protein being amenable to small molecule drug intervention. After seeding each cancer specific network with their respective drivers, we yielded a total of 631 distinct proteins across all cancers (**Supplementary Table 12**). The median number of unique proteins in each network across all cohorts was 57 with colorectal cancer possessing the largest network (n=231, **Fig. 4**) and spindle cell carcinoma (n=10) possessing the smallest network. As expected there was a correlation between network size and number of identified drivers for each cancer type (Pearson R = 0.9, *P*=1.23×10^-9^).

**FIgure 4.**
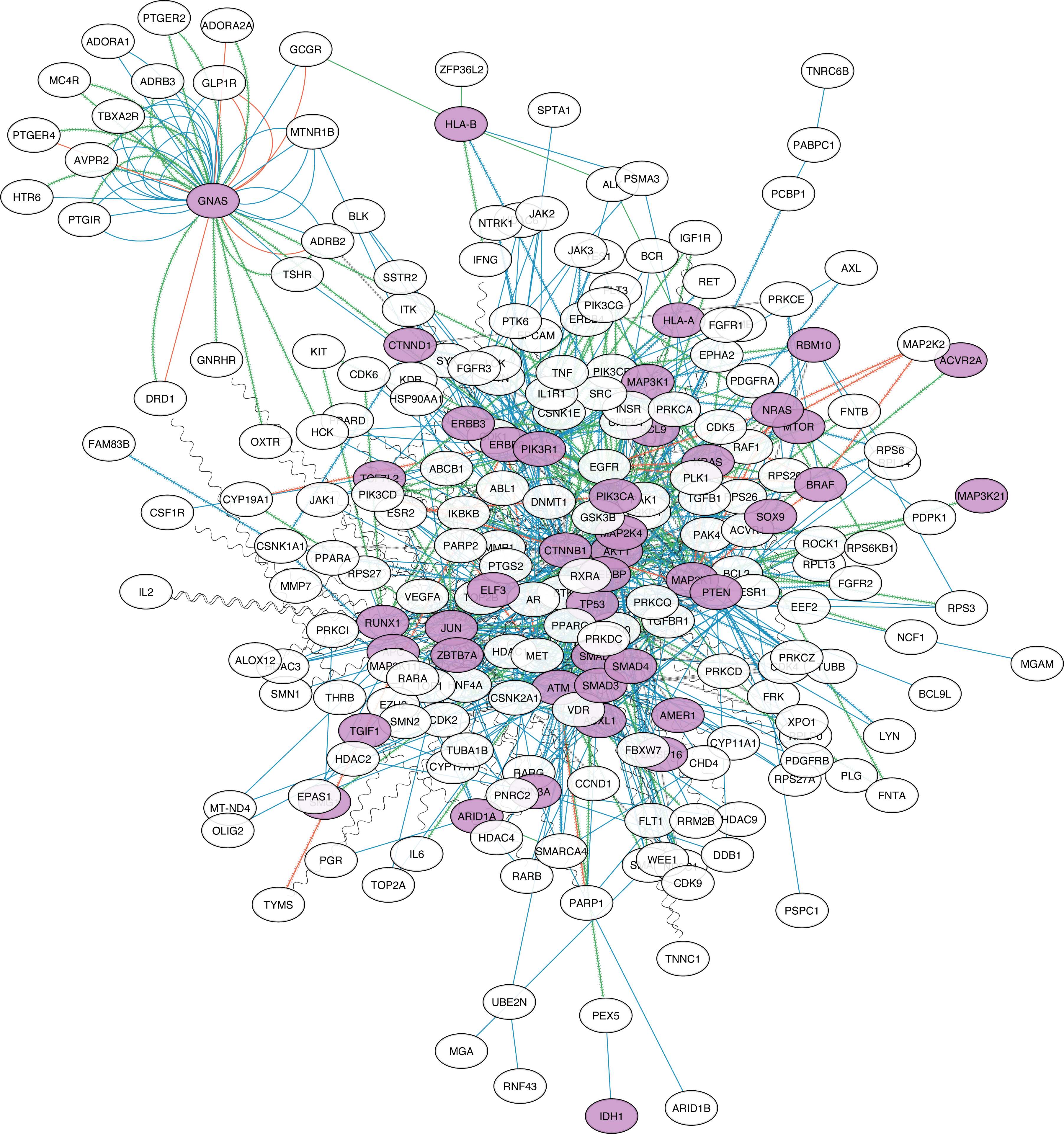
Example druggability networks for colorectal cancer. Nodes acting as cancer-specific drivers are shaded purple. Edge visual properties are as follows: OncoKB interactions, red contiguous arrow; Signor interactions, green contiguous arrow; Signor inhibitors, black vertical slash; complex, black zigzag; direct interaction, red solid line; direct X-ray interaction, green solid line; direct non-protein data bank interaction, blue solid line; reaction, blue contiguous arrow; transcriptional interaction, black sinewave.

Of these 631 proteins, 58% (n=369) were retrieved solely through network analysis of which the majority (n=323) were novel to any of the cancer types (hereon referred to as cancer-network proteins). Notable examples include *HDAC1*, *CDK2* and *CDK1* which were present in 31, 29 and 28 cohorts respectively. We observed 70% (n=225) of these cancer-network proteins as being targetable by existing approved or investigational therapies with notable examples including *BCL2* and *BTK*. Of the remaining 97 genes, 34 are commonly essential, 11 possess concordant lineage specificity, 48 are ligandable by 3D structure and 11 have an existing high quality probe available (**Supplementary Table 13**). Collectively these data provide potential future opportunities for therapy for a number of cancers.

## DISCUSSION

Delivering precision oncology to all patients is partly constrained because routine patient tumours are generally only testing for a restricted set of common actionable mutations. One of the main aims of the 100,000 Genomes Project is to improve cancer care for NHS patients through personalised medicine by implementing WGS as part of routine care^8^. Herein we have analysed WGS data on 10,470 patients recruited to the 100kGP study to explore the value of WGS to inform patient care. The strengths of this study not only include the cohort size but the combination of systematic processing of samples and data arising from multiple treatment centres across England.

Although we primarily focussed on point mutations and small indels we identified 330 cancer driver genes, 74 of which are novel to a cancer type. The similarities and differences in driver mutation frequencies in cancers arising from the same organ suggest shared and divergent pathways in oncogenesis. Importantly, many driver mutations are common across different tumour types. If clinically translated, these observations suggest currently 55% of patients harbour an actionable mutation, either in terms of predicting sensitivity to certain treatments or clinical trial eligibility. This contrasts to around 22% achievable if based on the routine small variant testing panels in widespread use^26^. While this is predicated on the assumption the use of approved drugs is a proxy for effective cancer therapies a recent study of cancer drug approvals by the U.S. Food and Drug Administration concluded that new cancer drug approvals reduce the risk of death and tumour progression^27^. To inform potential future therapeutic opportunities, we applied established chemogenomic technologies to map and pharmacologically annotate the cellular network of cancer genes identified by WGS. Through annotation of the cellular network with measures of essentiality and selectivity we are able to highlight additional potential therapeutic targets in cancer. It is likely that such endeavours will be improved upon through the use of high-throughput assays assessing more detailed functional consequences of somatic mutations^28^.

While the 100KGP was predicated on delivering diagnostic tests for well-established actionable mutations in NHS cancer patients with high sensitivity, concern has been raised of missing well-recognized clinically actionable mutations. In our analysis the frequency of established cancer-specific oncogenic drivers recovered was comparable to MSK-IMPACT and MSK-MET. Moreover, the sensitivity of 100x WGS to identify variants was high even for samples with low tumour purity (**Supplementary Methods**, **Supplementary Fig.7-11**). There are however technical limitations to current short read WGS, notably structural variants are not robustly identifiable with low concordance being a feature of currently implemented algorithms. While a consensus approach in the clinical setting can be adopted for the identification of such genomic features, it will likely only be addressed by adoption of long-read sequencing, albeit with a high requirement for DNA. Finally, the cost of WGS may be seen as currently prohibitive being around 12 times more costly than gene panels such as RMH200. Hence, as new panels informed by WGS findings are being developed, the attractiveness of WGS as a standalone test is questionable.

## Data Availability

Data is available through the Genomics England Platform.

## ACKNOWLEDGEMENTS

Funding was provided by the Wellcome Trust (214388), Cancer Research UK (C1298/A8362) and the Medical Research Council. A.S. is in receipt of a National Institute for Health Research (NIHR) Academic Clinical Lectureship, funding from the Royal Marsden Biomedical Research Centre, a starter grant for clinical lecturers from the Academy of Medical Sciences, and a Wellcome Trust Early Career Award (227000/Z/23/Z). This is a summary of independent research supported by the NIHR Biomedical Research Centre at the Royal Marsden NHS Foundation Trust and the Institute of Cancer Research.

This research was made possible through access to the data and findings generated by the 100,000 Genomes Project. The 100,000 Genomes Project is managed by Genomics England Limited (a wholly owned company of the Department of Health and Social Care). The 100,000 Genomes Project is funded by the National Institute for Health Research and NHS England. The Wellcome Trust, Cancer Research UK and the Medical Research Council also funded research infrastructure. The 100,000 Genomes Project uses data provided by patients and collected by the National Health Service as part of their care and support. We thank Alona Sosinsky, Scientific Director for Cancer at Genomics England, for the personal communication regarding the sensitivity of WGS for detection of well-established cancer driver mutations.

## GENOMICS ENGLAND RESEARCH CONSORTIUM

Ambrose, J. C.^1^; Arumugam, P.^1^; Bevers, R.^1^; Bleda, M.^1^; Boardman-Pretty, F. ^1,2^; Boustred, C. R.^1^; Brittain, H.^1^; Brown, M.A.^1^; Caulfield, M. J.^1,2^; Chan, G. C.^1^; Giess A.^1^; Griffin, J. N.^1^; Hamblin, A.^1^; Henderson, S.^1,2^; Hubbard, T. J. P. ^1^; Jackson, R.^1^; Jones, L. J.^1,2^; Kasperaviciute, D.^1,2^; Kayikci, M.^1^; Kousathanas, A.^1^; Lahnstein, L.^1^; Lakey, A.^1^; Leigh, S. E. A.^1^; Leong, I. U. S.^1^; Lopez, F. J.^1^; Maleady-Crowe, F.^1^; McEntagart, M.^1^; Minneci F.^1^; Mitchell, J.^1^; Moutsianas, L.^1,2^; Mueller, M.^1,2^; Murugaesu, N.^1^; Need, A. C.^1,2^; O‘Donovan P.^1^; Odhams, C. A.^1^; Patch, C.^1,2^; Perez-Gil, D.^1^; Pereira, M. B.^1^; Pullinger, J.^1^; Rahim, T.^1^; Rendon, A.^1^; Rogers, T.^1^; Savage, K.^1^; Sawant, K.^1^; Scott, R. H.^1^; Siddiq, A.^1^; Sieghart, A.^1^; Smith, S. C.^1^; Sosinsky, A.^1,2^; Stuckey, A.^1^; Tanguy M.^1^; Taylor Tavares, A. L.^1^; Thomas, E. R. A.^1,2^; Thompson, S. R.^1^; Tucci, A.^1,2^; Welland, M. J.^1^; Williams, E.^1^; Witkowska, K.^1,2^; Wood, S. M.^1,2^; Zarowiecki, M.^1^.

1. Genomics England, London, UK.
2. William Harvey Research Institute, Queen Mary University of London, London, UK.

## AUTHOR CONTRIBUTIONS

B.K, A.S, and R.S.H designed the study. B.K., A.S., A.C, D.C. performed sample curation, B.K., A.S., A.E., A.J.C., D.C., R.C., A.G., A.L., K.M., D.W., performed bioinformatic and statistical analysis. B.K. A.S., A.E. and R.S.H. drafted the manuscript; all authors reviewed, read, and approved the final manuscript.

## DATA AVAILABILITY

Summary statistics for each tumour group are provided in the supplementary tables where such data does not enable identification of participants. All sample-specific WGS data and processed files from the 100,000 Genomes Project can be accessed by joining the Pan Cancer Genomics England Clinical Interpretation Partnership (GeCIP) Domain once an individual’s data access has been approved (https://www.genomicsengland.co.uk/portfolio/pan-cancer-across-cancers-gecip315 domain/). The link to becoming a member of GECIP and having access can be found here https://www.genomicsengland.co.uk/research/academic/join-gecip. The process involves an online application, verification by the applicant’s institution, completion of a short information governance training course, and verification of approval by Genomics England. Please see https://www.genomicsengland.co.uk/research/academic for more information. The Genomics England data access agreement can be obtained from https://figshare.com/articles/GenomicEnglandProtocol_pdf/4530893/5. All analysis of Genomics England data must take place within the Genomics England Research Environment (https://www.genomicsengland.co.uk/understanding-genomics/data). The 100,000 Genomes Project publication policies can be obtained from https://www.genomicsengland.co.uk/about-gecip/publications. Samples and results used in this study are provided in Genomics England under /re_gecip/shared_allGeCIPs/pancancer_drivers/results/. The COSMIC and OncoKB clinical actionability data are available from https://cancer.sanger.ac.uk/actionability and https://www.oncokb.org/actionableGenes#sections=Tx, respectively. The canSAR chemogenomics data are available from https://cansar.ai/. The NHS Genomic Test Directory for Cancer is available from https://www.england.nhs.uk/publication/national-genomic-test-directories/.

## CODE AVAILABILITY

Details and code for using the Intogen framework are available here (https://intogen.readthedocs.io/en/latest/index.html). The specific code to perform this analysis is available in the Genomics England research environment under /re_gecip/shared_allGeCIPs/pancancer_drivers/code/. Code will be made publicly available in a dedicated Github repository on publication of the manuscript. The code to perform the canSAR chemogenomics analysis is available through Zenodo (https://zenodo.org/record/8329054).

## REFERENCES

1. Topol, E. J. Individualized medicine from prewomb to tomb. Cell 157, 241–253 (2014).

2. Spear, B. B., Heath-Chiozzi, M. & Huff, J. Clinical application of pharmacogenetics. Trends in Molecular Medicine vol. 7 201–204 Preprint at 10.1016/s1471-4914(01)01986-4 (2001).

3. Maeda, H. & Khatami, M. Analyses of repeated failures in cancer therapy for solid tumors: poor tumor-selective drug delivery, low therapeutic efficacy and unsustainable costs. Clin. Transl. Med. 7, 11 (2018).

4. Schwartzberg, L., Kim, E. S., Liu, D. & Schrag, D. Precision Oncology: Who, How, What, When, and When Not? American Society of Clinical Oncology Educational Book 160–169 Preprint at 10.1200/edbk_174176 (2017).

5. Stratton, M. R., Campbell, P. J. & Andrew Futreal, P. The cancer genome. Nature vol. 458 719–724 Preprint at 10.1038/nature07943 (2009).

6. Chakravarty, D., et al. OncoKB: A Precision Oncology Knowledge Base. JCO Precis Oncol 2017, (2017).

7. Freedman, A. N., et al. Use of Next-Generation Sequencing Tests to Guide Cancer Treatment: Results From a Nationally Representative Survey of Oncologists in the United States. JCO Precis Oncol 2, (2018).

8. Turnbull, C. et al. The 100 000 Genomes Project: bringing whole genome sequencing to the NHS. BMJ 361, k1687 (2018).

9. Martínez-Jiménez, F. et al. A compendium of mutational cancer driver genes. Nat. Rev. Cancer 20, 555–572 (2020).

10. Tate, J. G. et al. COSMIC: the Catalogue Of Somatic Mutations In Cancer. Nucleic Acids Res. 47, D941–D947 (2019).

11. Nguyen, B. et al. Genomic characterization of metastatic patterns from prospective clinical sequencing of 25,000 patients. Cell 185, 563–575.e11 (2022).

12. Cheng, D. T. et al. Memorial Sloan Kettering-Integrated Mutation Profiling of Actionable Cancer Targets (MSK-IMPACT): A Hybridization Capture-Based Next-Generation Sequencing Clinical Assay for Solid Tumor Molecular Oncology. J. Mol. Diagn. 17, 251–264 (2015).

13. Salipante, S. J., Scroggins, S. M., Hampel, H. L., Turner, E. H. & Pritchard, C. C. Microsatellite Instability Detection by Next Generation Sequencing. Clinical Chemistry vol. 60 1192–1199 Preprint at 10.1373/clinchem.2014.223677 (2014).

14. Davies, H. et al. HRDetect is a predictor of BRCA1 and BRCA2 deficiency based on mutational signatures. Nat. Med. 23, 517–525 (2017).

15. Mitsopoulos, C. et al. canSAR: update to the cancer translational research and drug discovery knowledgebase. Nucleic Acids Res. 49, D1074–D1082 (2020).

16. Orchard, S. et al. The MIntAct project--IntAct as a common curation platform for 11 molecular interaction databases. Nucleic Acids Res. 42, D358–63 (2014).

17. Hornbeck, P. V., et al. PhosphoSitePlus, 2014: mutations, PTMs and recalibrations. Nucleic Acids Res. 43, D512–20 (2015).

18. Shimada, K., Bachman, J. A., Muhlich, J. L. & Mitchison, T. J. shinyDepMap, a tool to identify targetable cancer genes and their functional connections from Cancer Dependency Map data. Elife 10, (2021).

19. ICGC/TCGA Pan-Cancer Analysis of Whole Genomes Consortium. Pan-cancer analysis of whole genomes. Nature 578, 82–93 (2020).

20. Bailey, M. H. et al. Comprehensive Characterization of Cancer Driver Genes and Mutations. Cell 173, 371–385.e18 (2018).

21. André, F. et al. Alpelisib for PIK3CA-Mutated, Hormone Receptor-Positive Advanced Breast Cancer. N. Engl. J. Med. 380, 1929–1940 (2019).

22. Fell, J. B. et al. Identification of the Clinical Development Candidate, a Covalent KRAS Inhibitor for the Treatment of Cancer. J. Med. Chem. 63, 6679–6693 (2020).

23. Canon, J. et al. The clinical KRAS(G12C) inhibitor AMG 510 drives anti-tumour immunity. Nature 575, 217–223 (2019).

24. Mateo, J. et al. A First-Time-in-Human Study of GSK2636771, a Phosphoinositide 3 Kinase Beta-Selective Inhibitor, in Patients with Advanced Solid Tumors. Clin. Cancer Res. 23, 5981–5992 (2017).

25. Marabelle, A. et al. Association of tumour mutational burden with outcomes in patients with advanced solid tumours treated with pembrolizumab: prospective biomarker analysis of the multicohort, open-label, phase 2 KEYNOTE-158 study. Lancet Oncol. 21, 1353–1365 (2020).

26. Website. ‘National Genomic Test Directory.’ n.d. https://www.england.nhs.uk/publication/national-genomic-test-directories/.

27. Michaeli, D. T. & Michaeli, T. Overall Survival, Progression-Free Survival, and Tumor Response Benefit Supporting Initial US Food and Drug Administration Approval and Indication Extension of New Cancer Drugs, 2003-2021. J. Clin. Oncol. 40, 4095–4106 (2022).

28. Kim, Y., et al. High-throughput functional evaluation of human cancer-associated mutations using base editors. Nat. Biotechnol. 40, 874–884 (2022).

