## Supplementary Fig. for "Cancer driver genes and opportunities for precision oncology revealed by whole genome sequencing 10,478 cancers"

Supplementary Figures

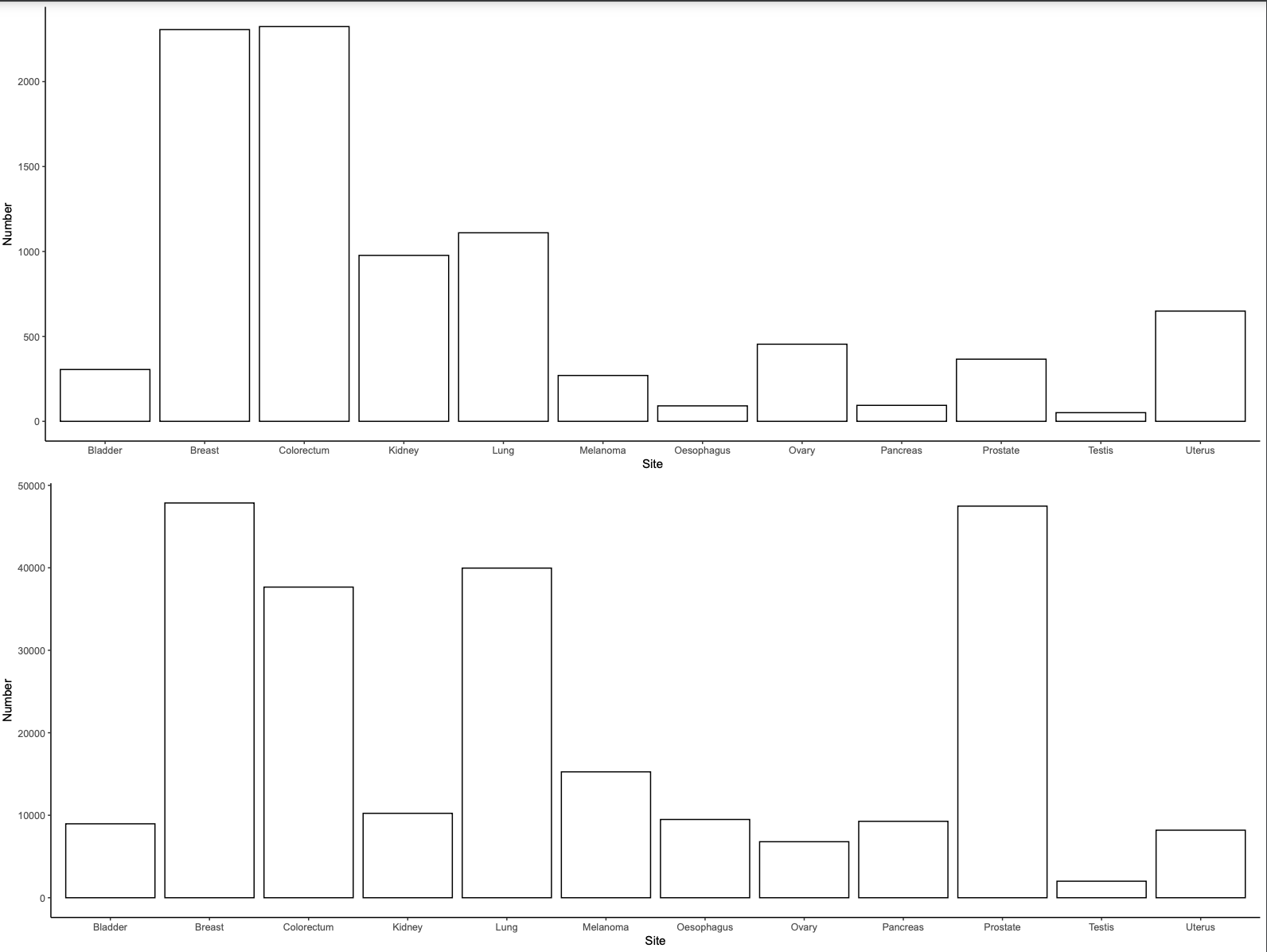

**Supplementary Figure 1. Comparison of number of samples per tumour type in the pan-cancer cohort compared to all cancer diagnosed in England in 2019.**

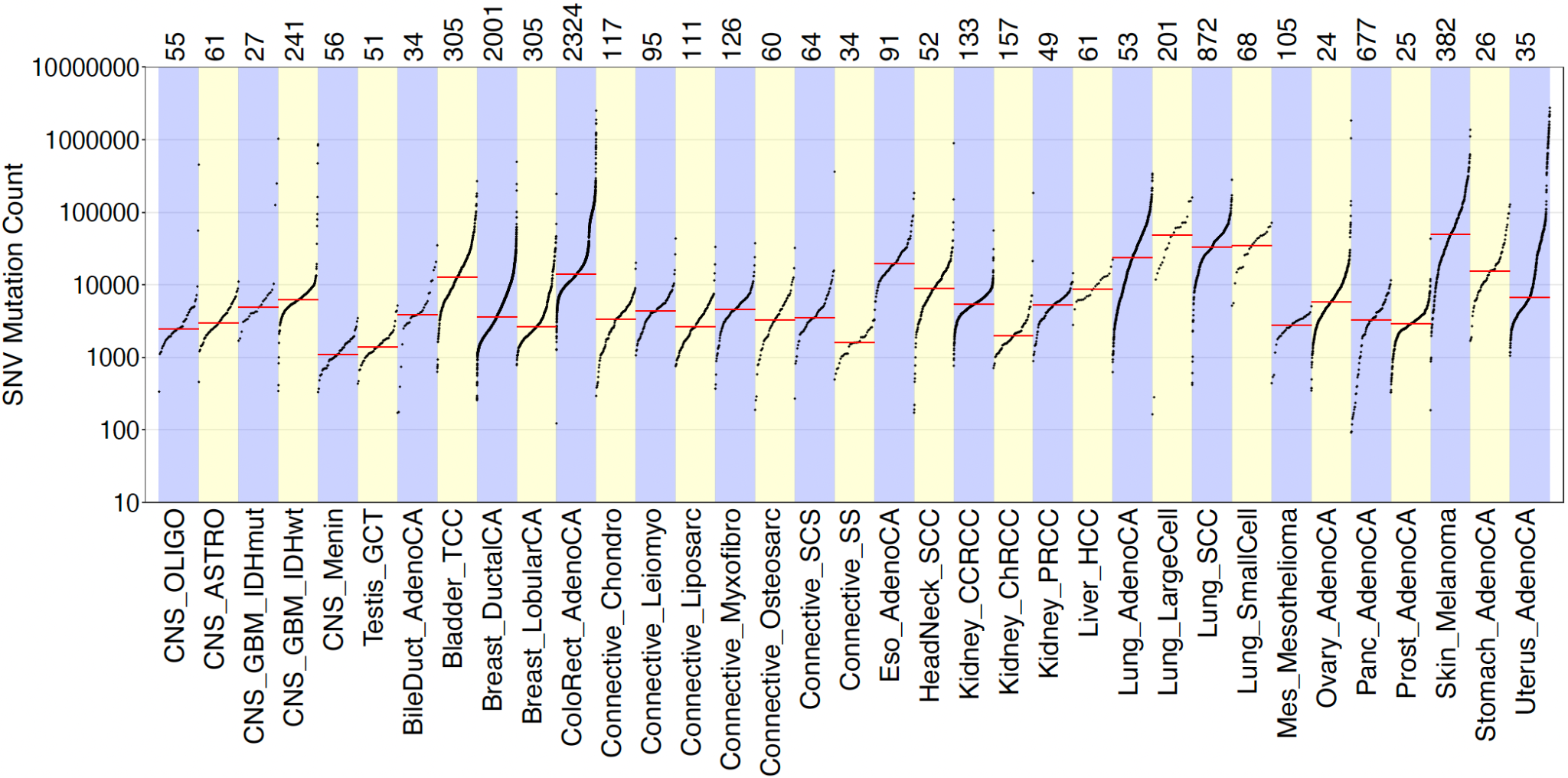

**Supplementary Figure 2. Mutation burden of tumours from each tumour type.** The number of samples contributing to each tumour type are shown above the plot. SNV, single nucleotide variant.

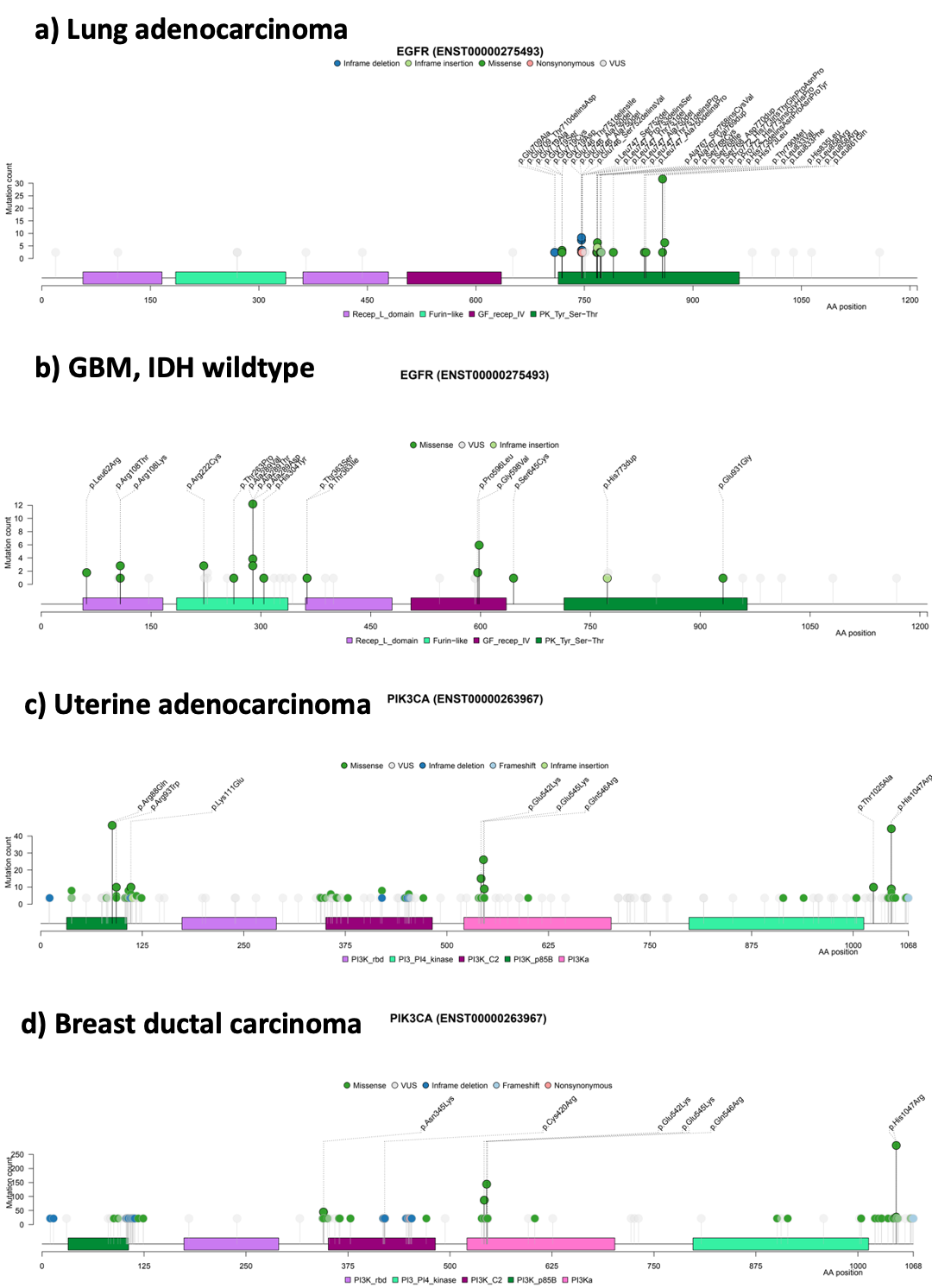

**Supplementary Figure 3: Driver mutation plots and pfam domain overlap.** a) Lung adenocarcinoma in EGFR; b) GBM IDH wildtype in EGFR; c) Uterine adenocarcinoma in PIK3CA; d) Breast ductal carcinoma in PIK3CA

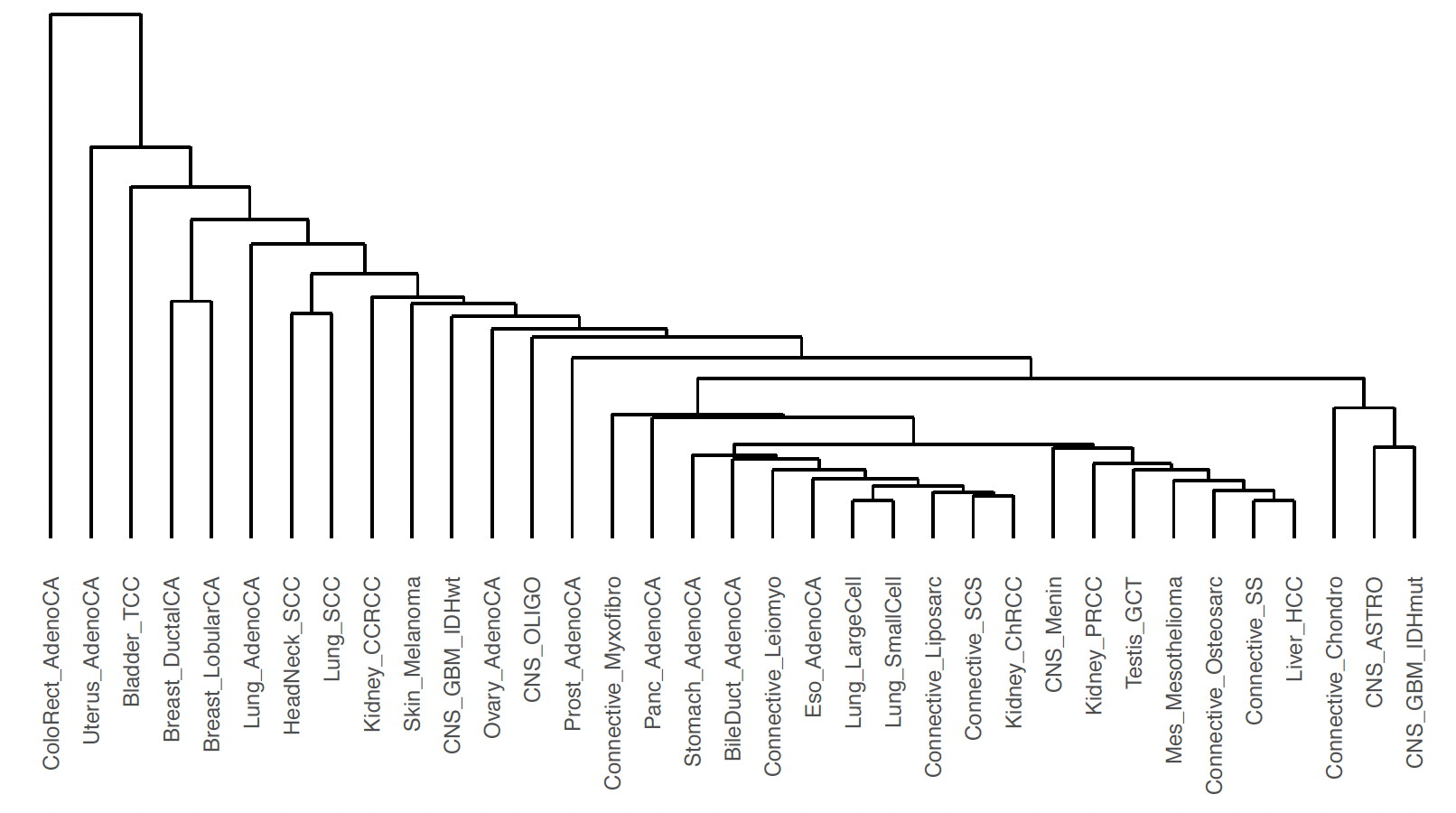

**Supplementary Figure 4. Hierarchical clustering of tumour types based on *P*-value of driver genes across the 35 cohorts.**

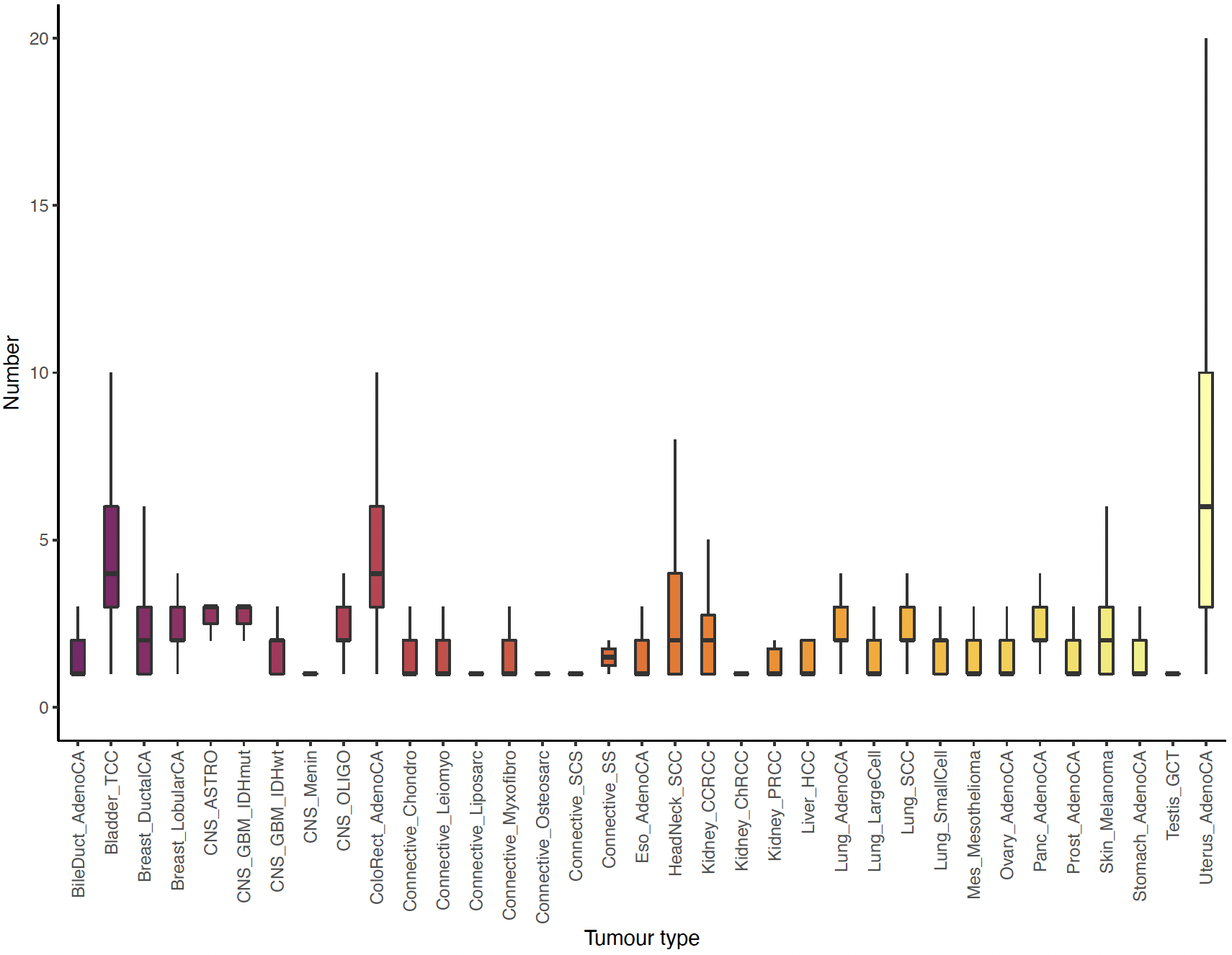

**Supplementary Figure 5. Per-tumour distribution of oncogenic mutations in tumour specific cancer driver genes, across the 35 cancer types.** Analysis restricted to driver genes as predicted by IntOGen in the given cancer type. Oncogenicity predicted using OncoKB.

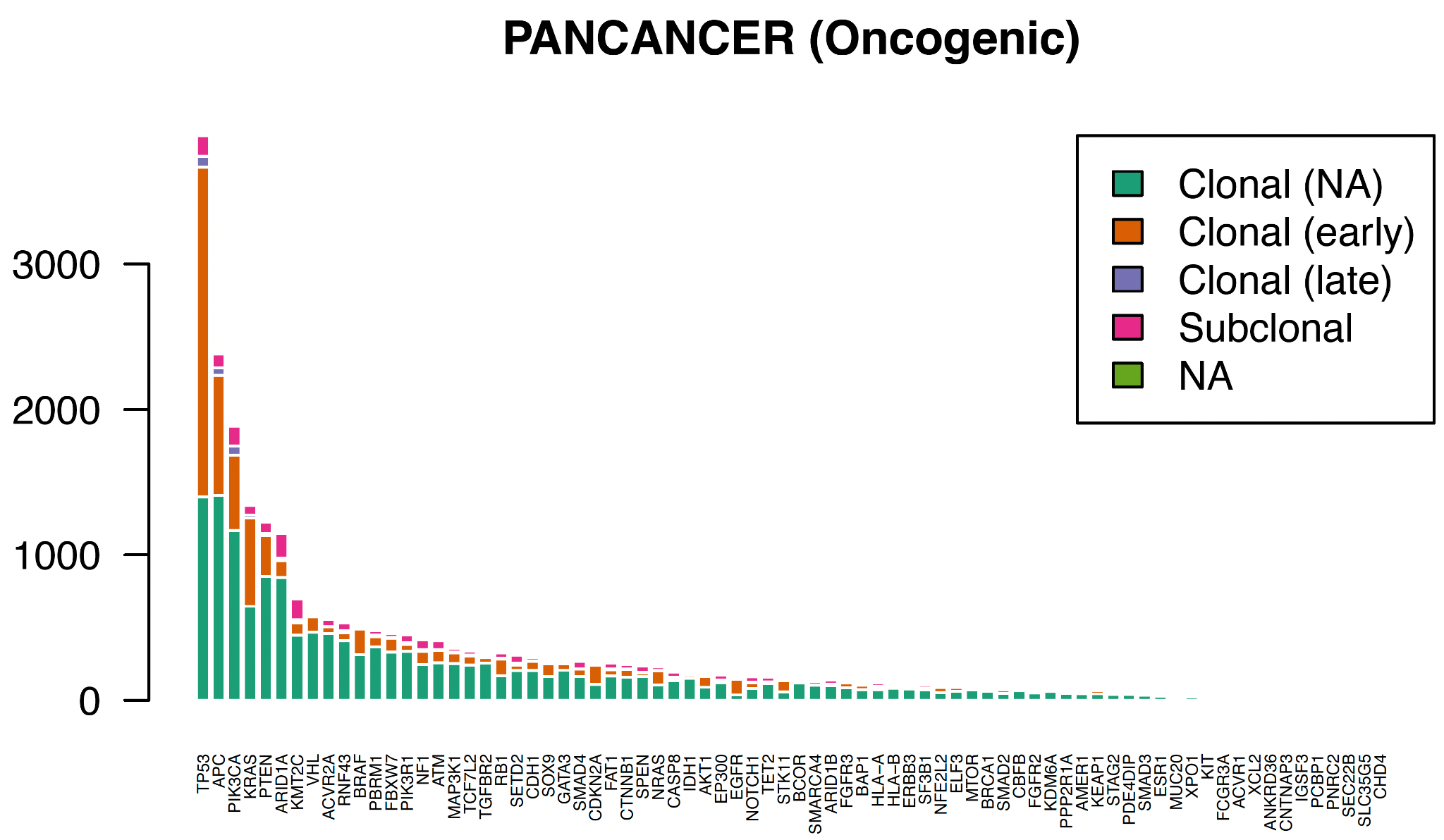

**Supplementary Figure 6: Oncogenic clonal and subclonal mutations across candidate driver genes pancancer.**

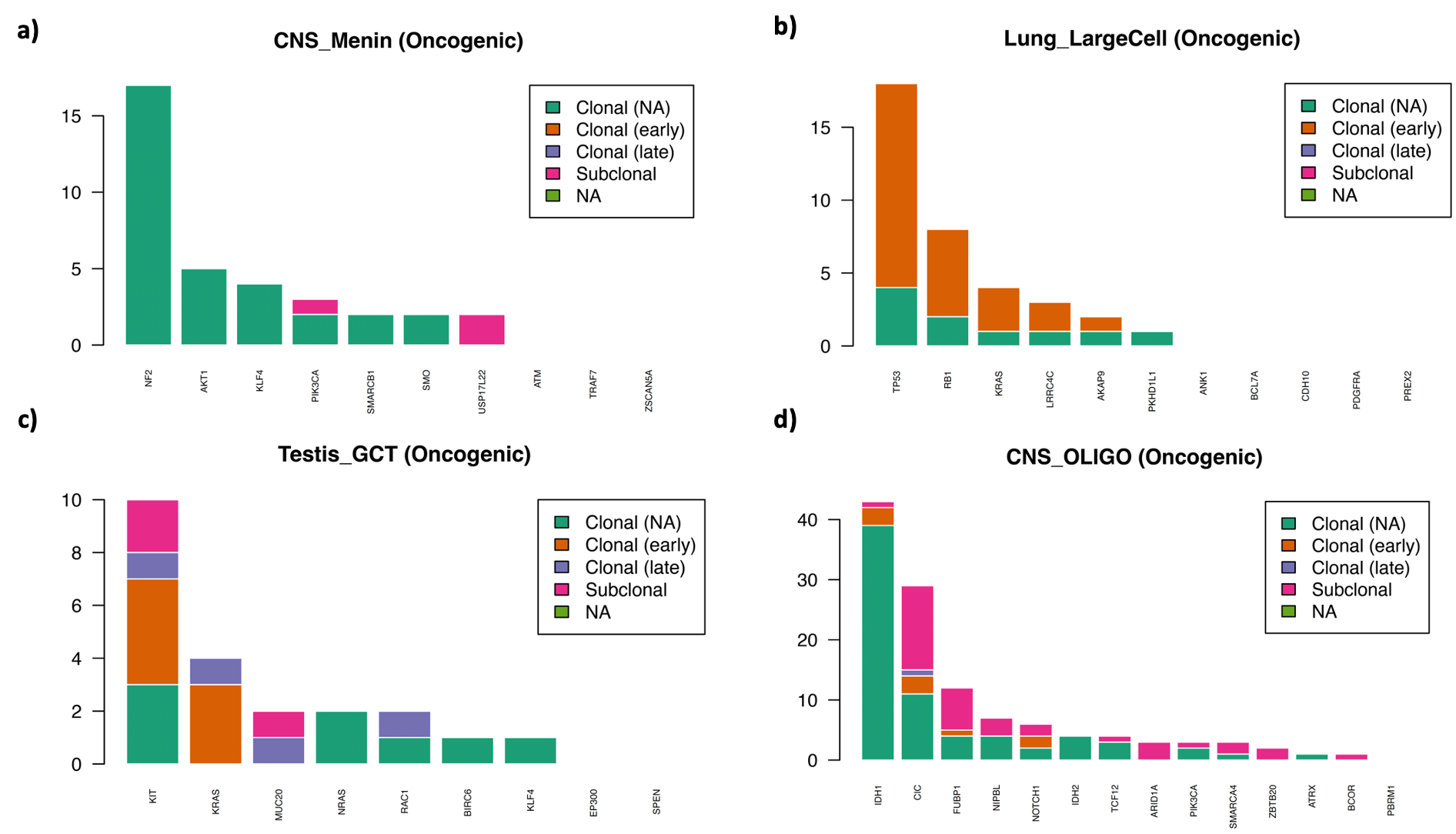

**Supplementary Figure 7: Oncogenic clonal and subclonal mutations across candidate driver genes in: a) Meningioma; b) Large Cell lung cancer; c) Testicular germ cell tumour; d) Oligodendroglioma.**

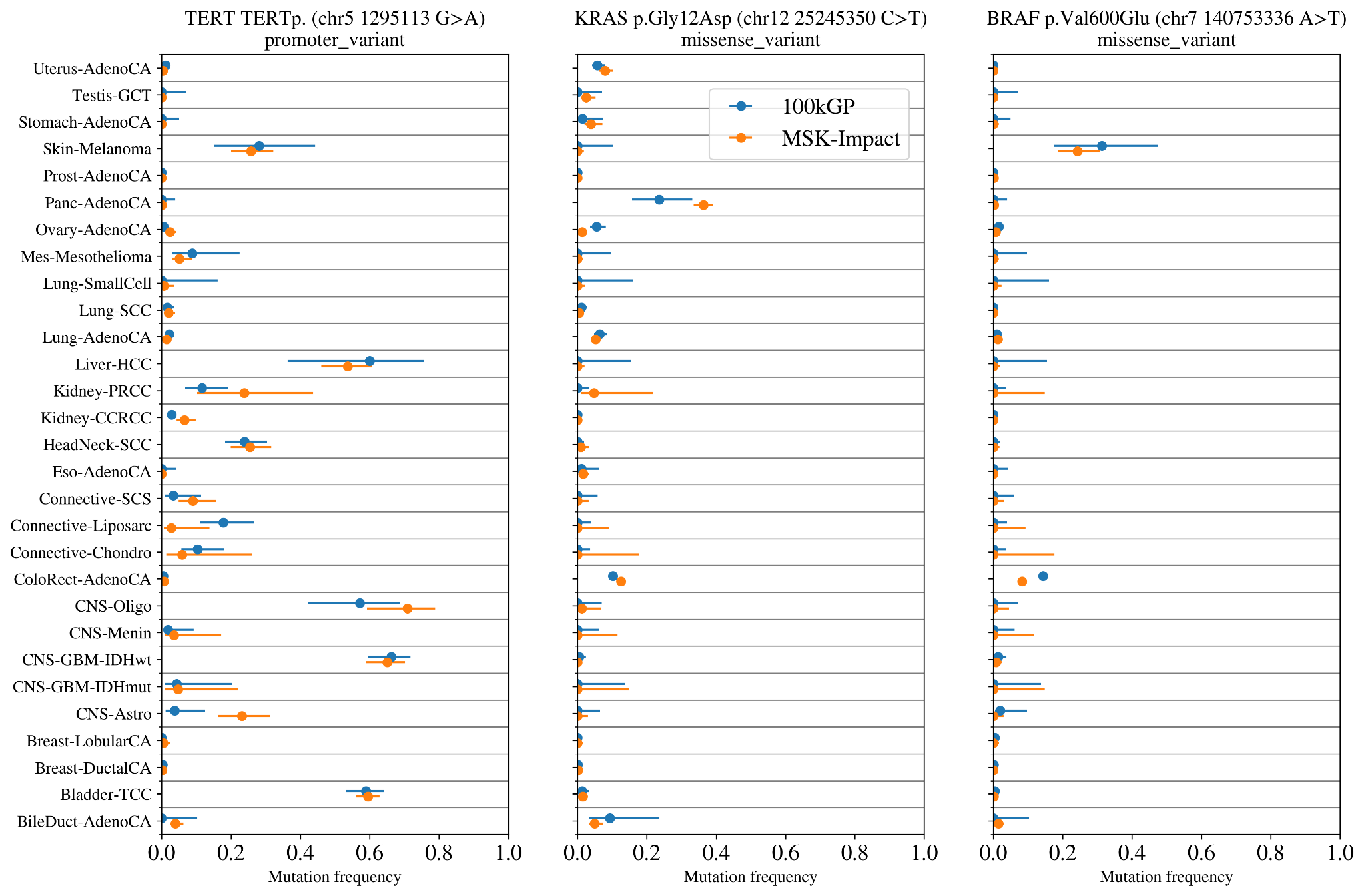

**Supplementary Figure 8. The rate of hotspot mutations in the 100kGP sample compared with panel sequencing in MSK IMPACT study.** The fraction of the samples with the hotspot mutation with 95% confidence intervals (binomial distribution with uniform prior).

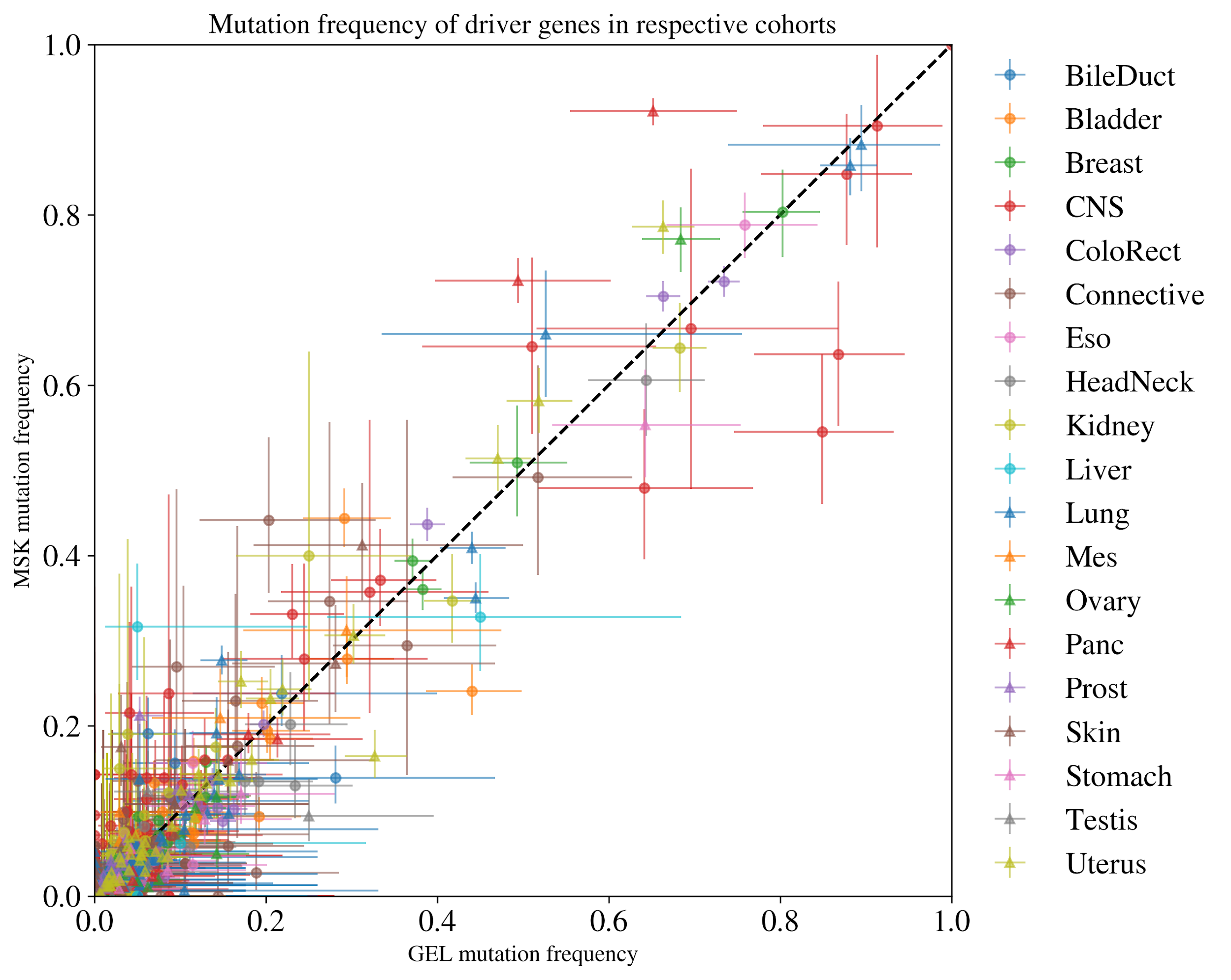

**Supplementary Figure 9. The rate of mutations in driver genes in the100kGP cohort compared to the MSK cohort.** The fraction of the samples with a hotspot mutation in the tumour group (95% confidence intervals for a binomial distribution with uniform prior). Each point corresponds to one of the 770 driver genes in tumour groups where the colour and point shape corresponds to the organ of the tumour group.

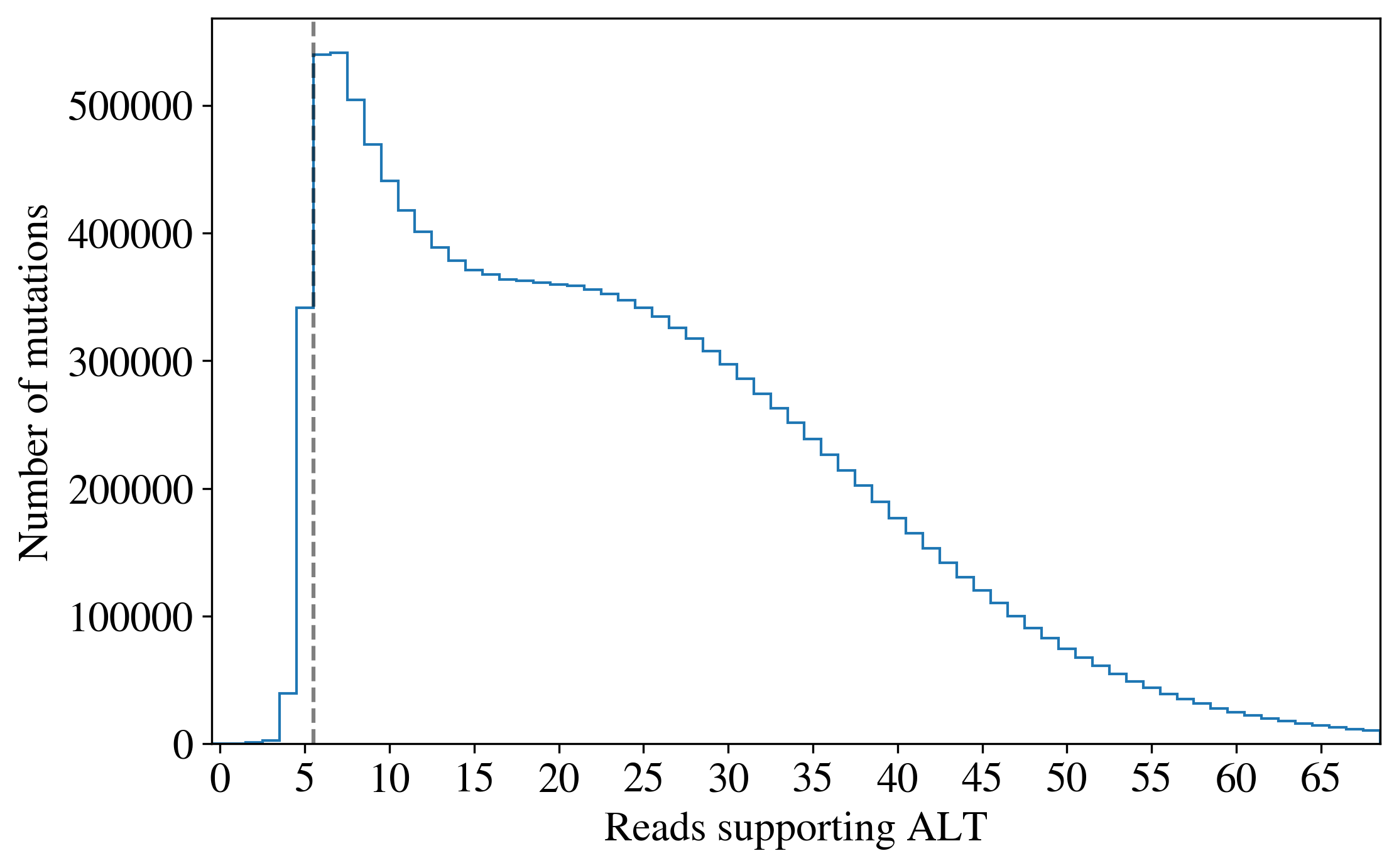

**Supplementary Figure 10. Distribution of read counts supporting the alternate allele across all PASS mutations in all tumour samples.** The steep dropoff in called mutations with fewer than 6 reads supporting the alternate allele suggests that a threshold of 6 is a reasonable proxy for selection criteria of Strelka variant calls.

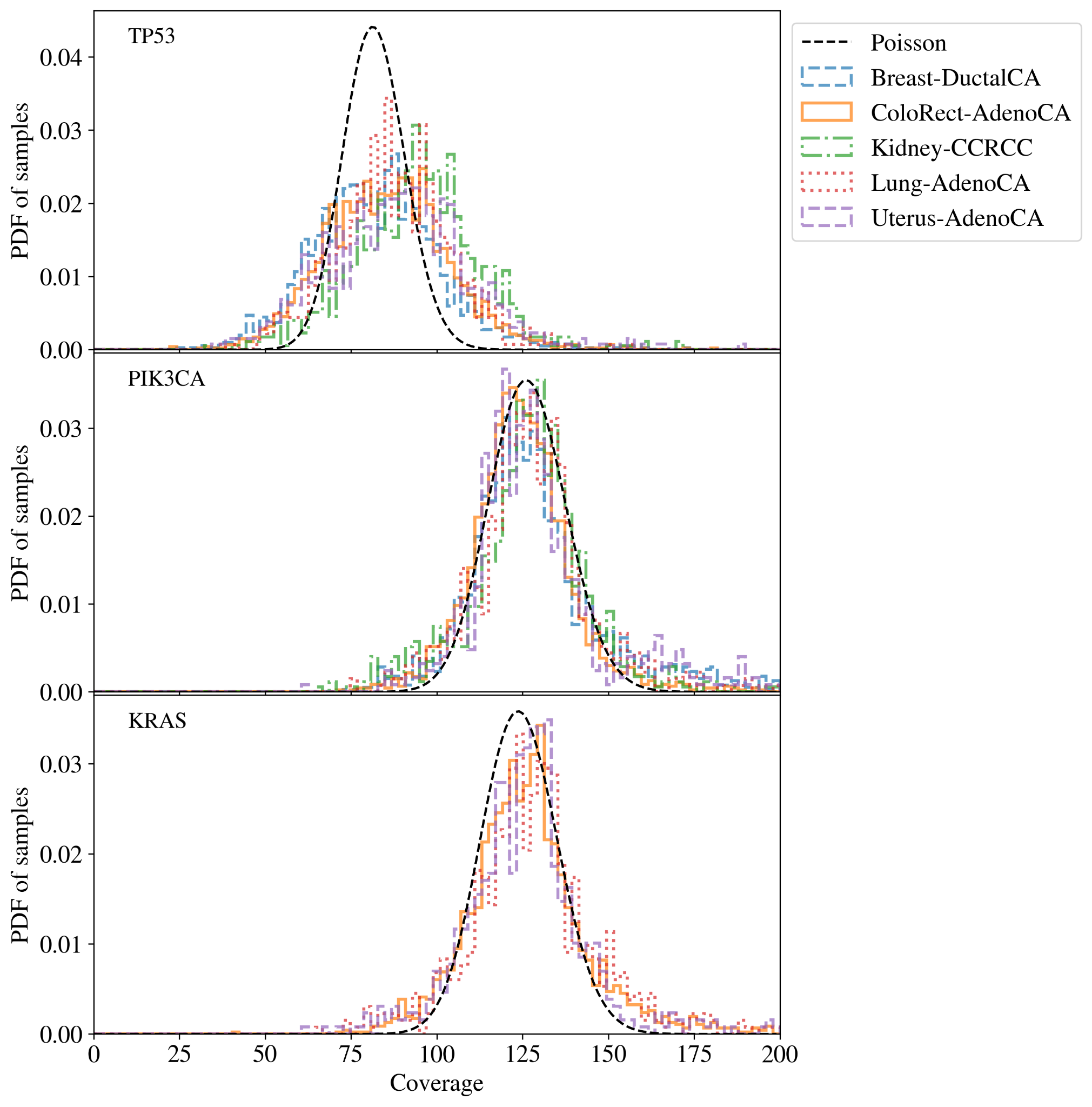

**Supplementary Figure 11. The distribution average read coverage for each sample in the given tumour group for the given gene.** Most genes such as PIK3CA and KRAS have greater than 100x coverage in the majority of samples and spread consistent with random Poisson noise. TP53 has significantly lower and more varied coverage between samples.

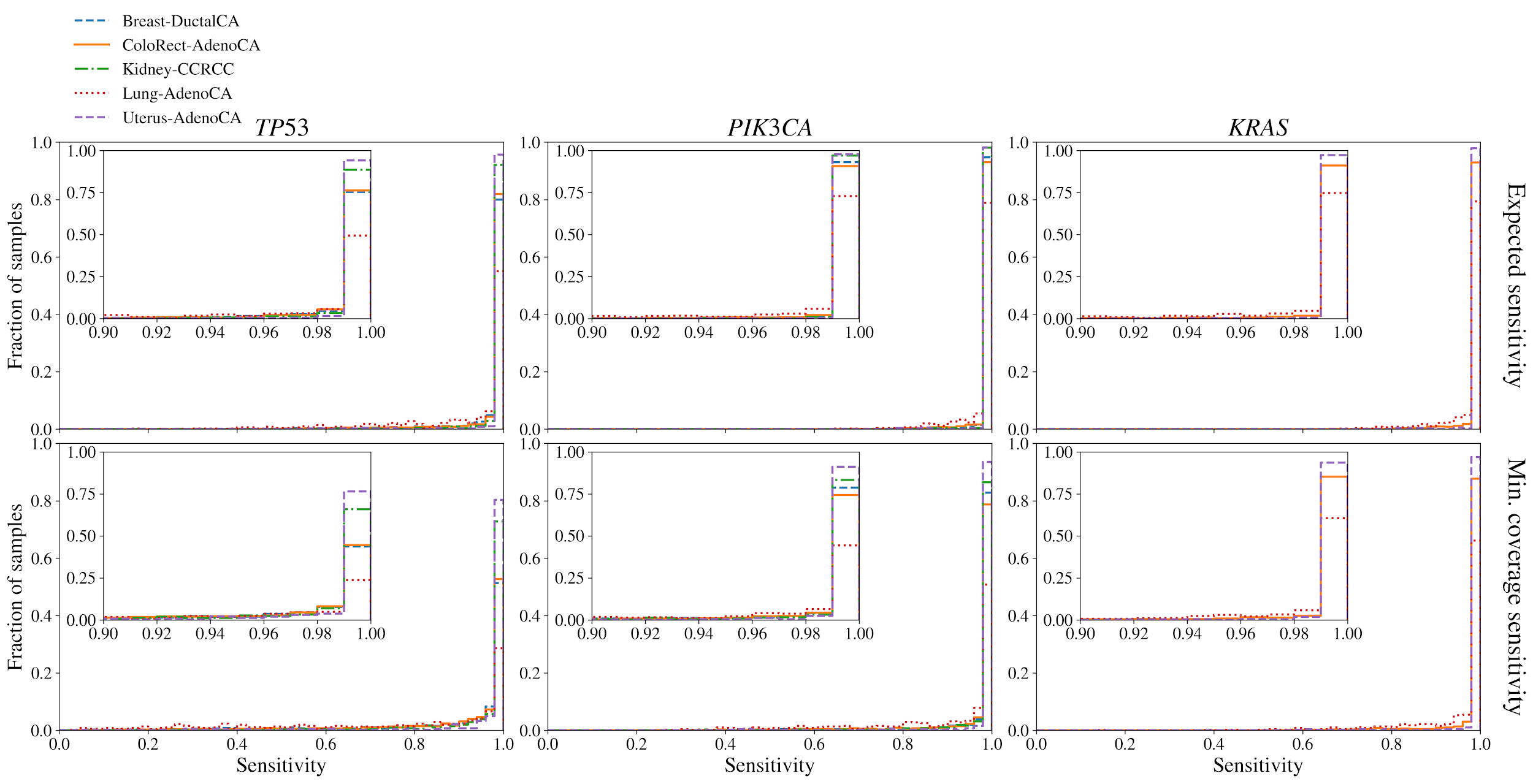

**Supplementary Figure 12. Distribution of mean sensitivity (top) and minimum sensitivity (bottom) across samples in the given tumour group for the given gene.** The mean sensitivity is estimated as the Poisson probability that there will be at least 6 alternate allele reads given the purity and mean gene coverage for the sample while the minimum sensitivity is given the minimum coverage across the gene. Across 88% of genes and samples have greater than 99% mean sensitivity.

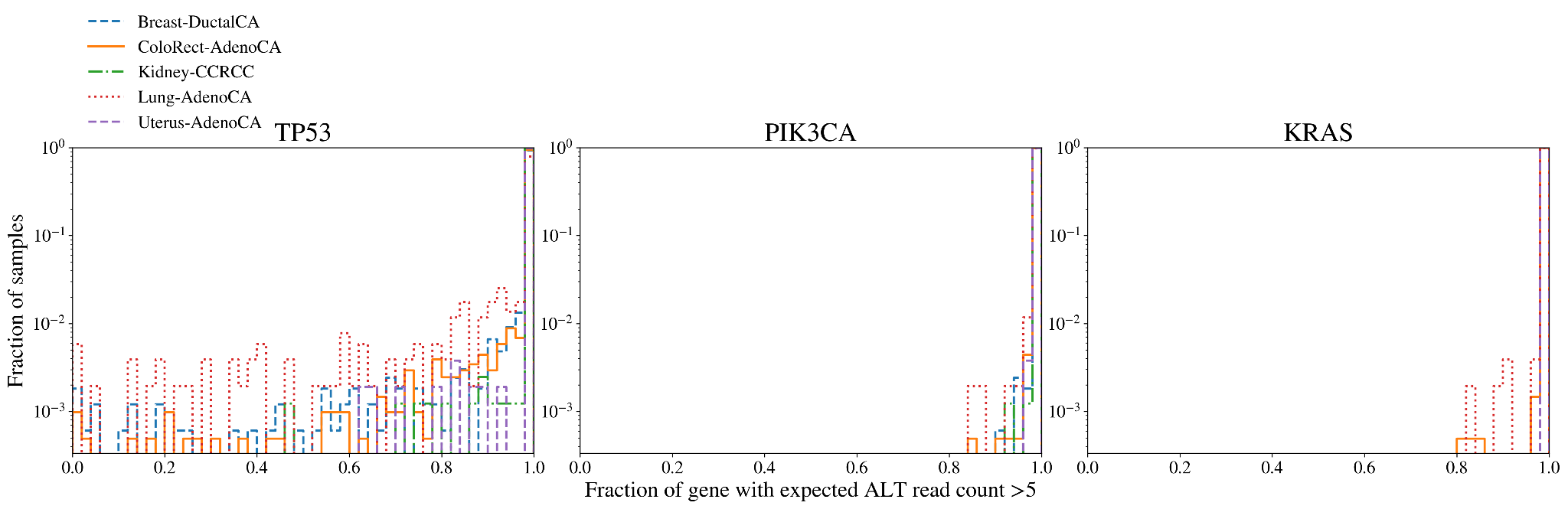

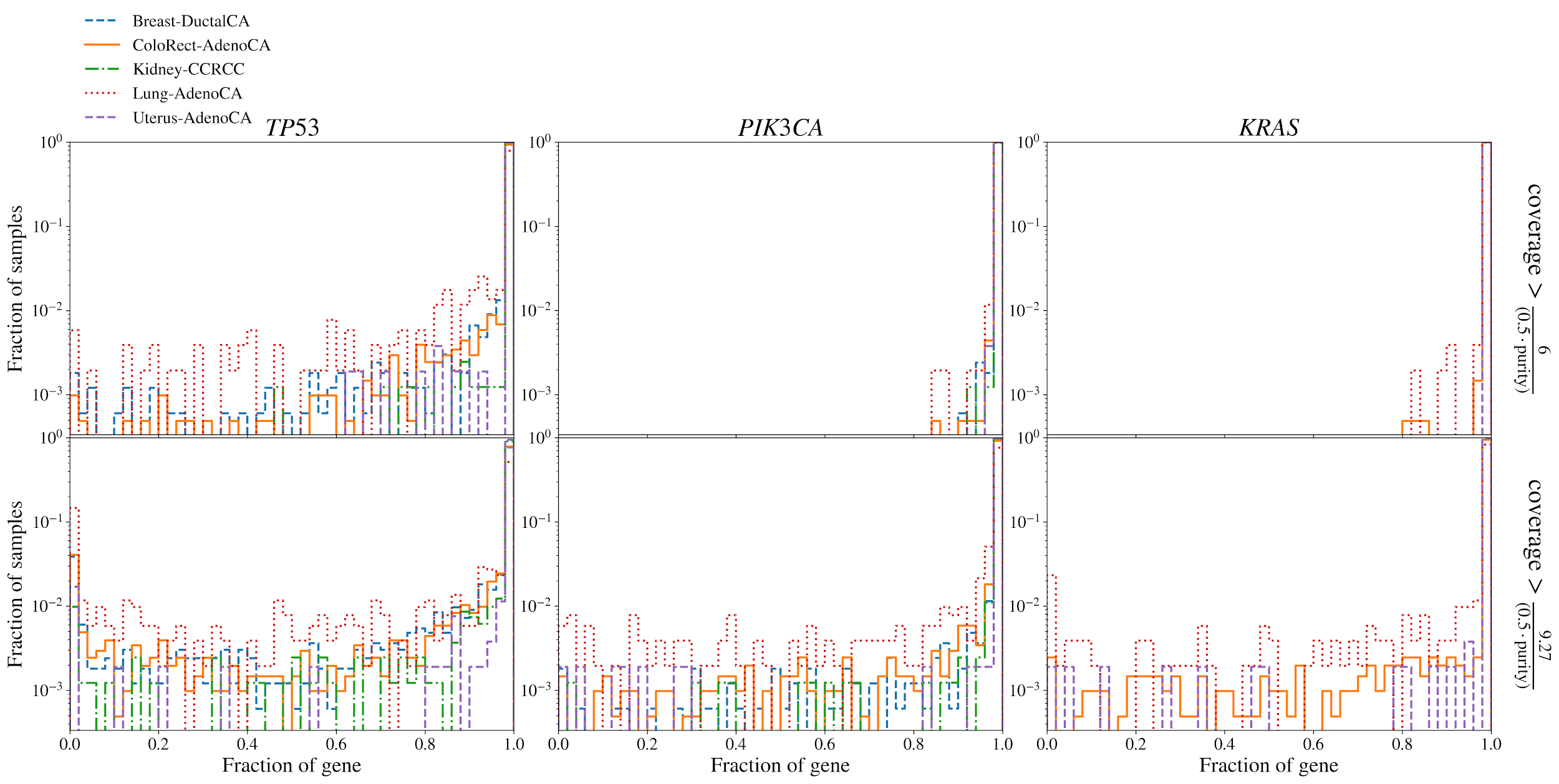

**Supplementary Figure 13. Fraction of gene where the coverage is high enough that the sensitivity would be at least 50% (top) and 90% (bottom).** The expected read count supporting alternate alleles is given by $0.5 \times coverage \times purity$. 6 alternate reads would give approximately a 50% chance of detecting a variant and 9.27 reads corresponds to approximately 90%. In 90% of all genes and samples, over 98% of the gene has sufficient coverage and purity to have an expected alternate read count greater than 6.

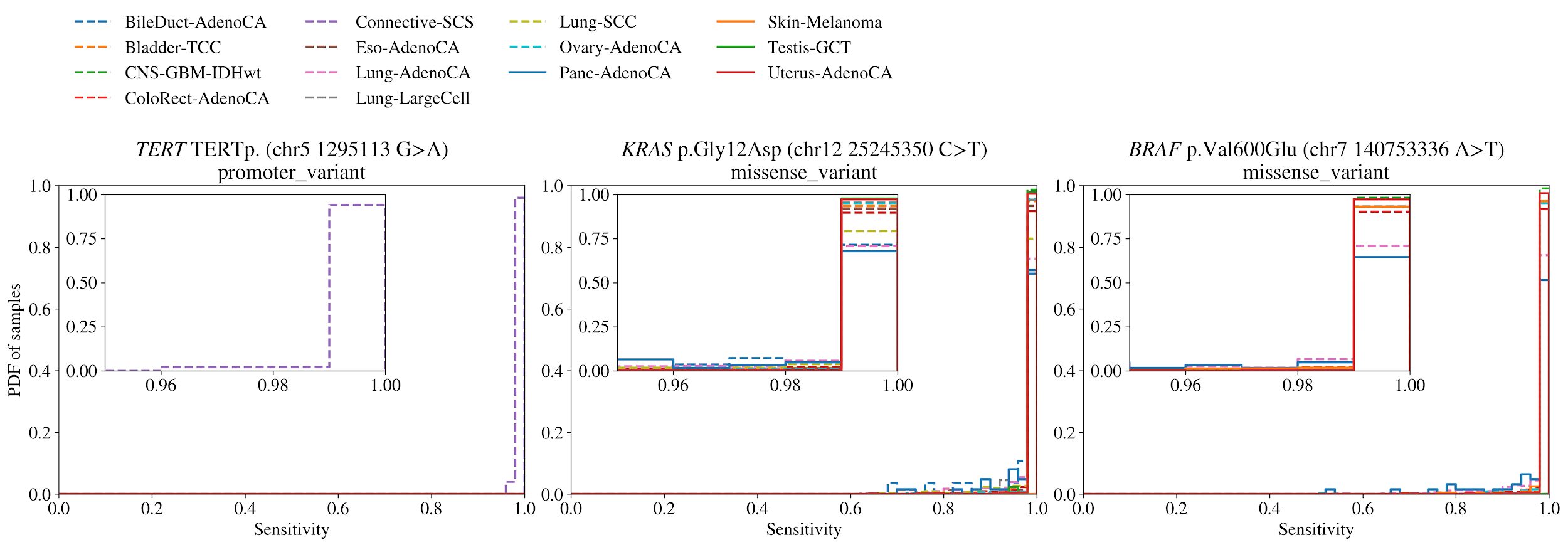

**Supplementary Figure 14. The distribution of expected sensitivity to hotspot mutations across samples in the given tumour group.** The sensitivity is estimated based on the Poisson probability of there being at least 6 alternate allele reads given the coverage and sample purity. 88% of hotspots in samples have greater than 99% sensitivity.

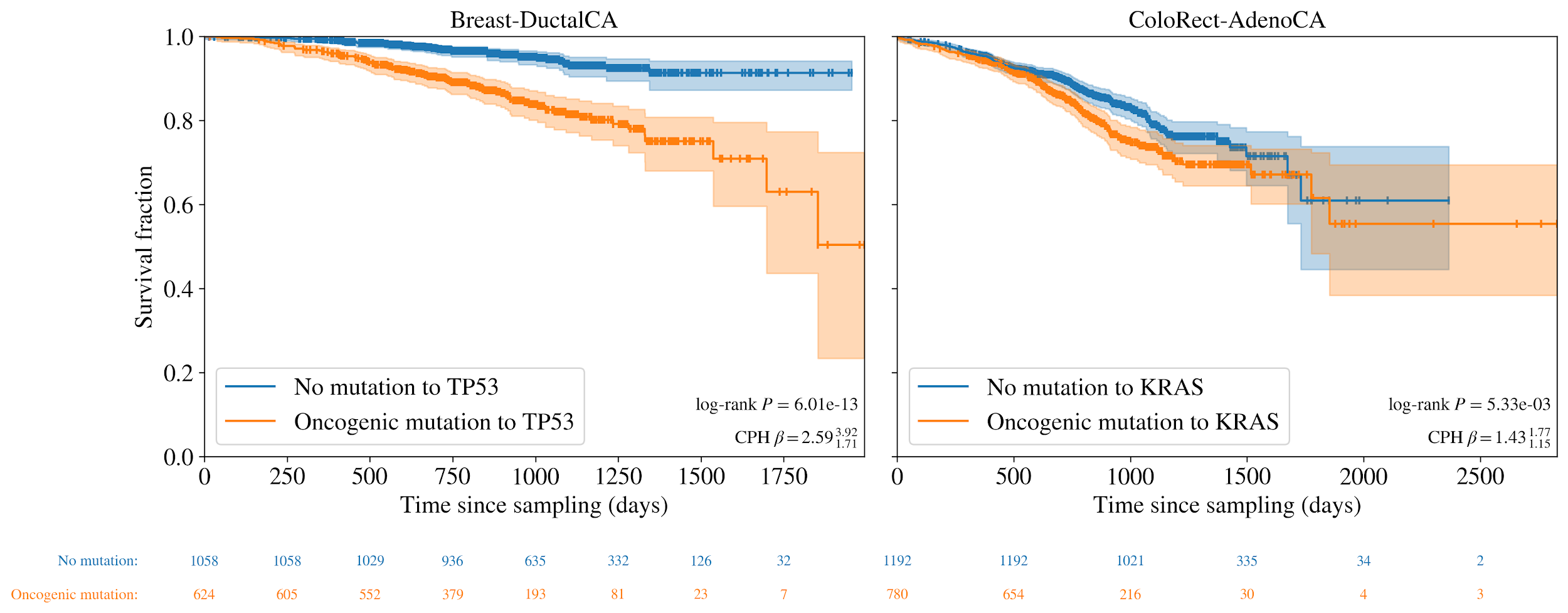

**Supplementary Figure 15. Survival curves for Breast-DuctalCA and ColoRect-AdenoCA cohorts with oncogenic mutations to TP53 and KRAS respectively vs those without.** Log-rank *P*values are shown in the bottom right along with coefficients from Cox Proportional Hazard (CPH) tests controlling for age, sex and principal components of population.

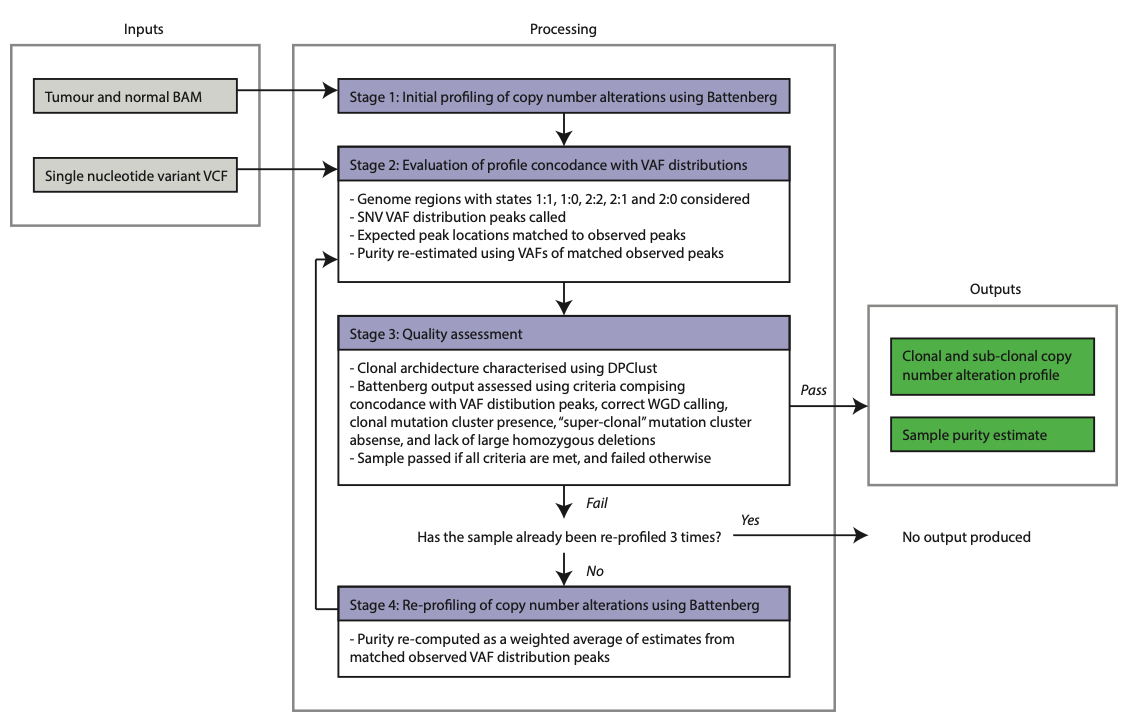

**Supplementary Figure 16: CNV calling pipeline overview.** BAM, binary sequence alignment map; VCF, variance call format; SNV, single nucleotide variant; VAF, variance allele frequency; WGD, whole genome duplication.

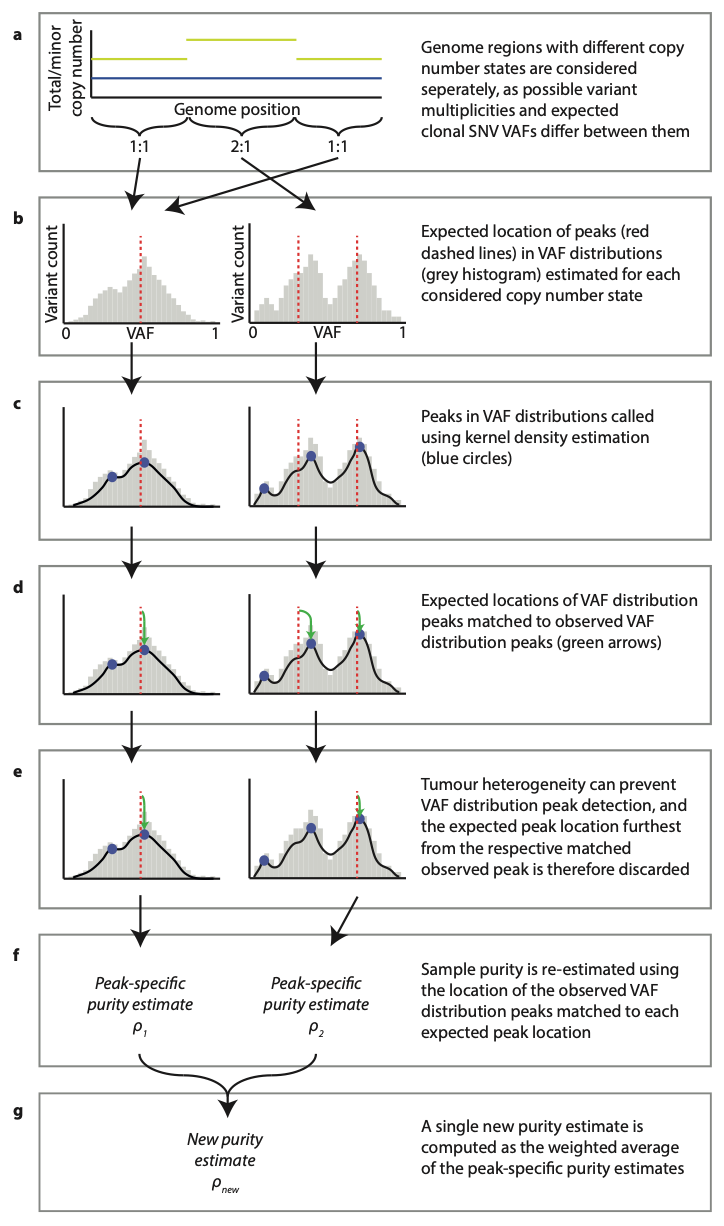

**Supplementary Figure 17: Overview of stage 2 of the CNV calling pipeline.** SNV, single nucleotide variant; VAF, variant allele frequency.
