## Supplementary Methods for "Cancer driver genes and opportunities for precision oncology revealed by whole genome sequencing 10,478 cancers"

**Curation of clinical data**

Clinical data were obtained from Public Health England’s National Cancer Registration and Analysis Service (PHE-NCRAS), NHS Digital (NHSD) and the Genomic Medicine Centres (GMCs) via the Genomics England Research Environment. Additional data were obtained from histopathology reports. Sequenced samples were matched to their respective PHE-NCRAS records using the date of tumour sampling and the PHE-NCRAS treatment dates, allowing maximum discrepancies of 28 days. Collected data comprised sex, year of birth, date of cancer diagnosis, date of last reported clinical follow-up, survival outcome with date of death if relevant, whether the sample was taken from a primary tumour, a metastasis or a recurrence of a primary tumour, and tumour histology. For some variables, data were obtained from multiple sources (*e.g*., GMC, NHSD, PHE-NCRAS). Potential conflicts between data sources were resolved by manual review. Sequenced tumours were assigned to one of 35 tumour groups based on originating tissue and histology: cholangiocarcinoma, bladder cancer, breast ductal carcinoma, breast lobular carcinoma, astrocytoma, glioblastoma (IDH mutated and wild type), meningioma, oligodendroglioma, colorectal adenocarcinoma, chondrosarcoma, leiomyosarcoma, liposarcoma, myxofibrosarcoma, osteosarcoma, spindle cell sarcoma, synovial sarcoma, esophageal adenocarcinoma, squamous cell carcinoma of the head and neck, clear cell renal cell carcinoma, chromophobe renal cell carcinoma, papillary renal cell carcinoma, hepatocellular carcinoma, lung adenocarcinoma, large-cell lung cancer, squamous cell carcinoma of the lung, small cell carcinoma of the lung, mesothelioma, ovarian adenocarcinoma, pancreatic adenocarcinoma, prostate adenocarcinoma, melanoma of the skin, gastric adenocarcinoma, testicular germ cell tumour and uterine adenocarcinoma (**Supplementary Table 1** and **2**).

NHSD and PHE-NCRAS data were used to identify participants that had received systemic treatment or sequenced-cancer-associated radiotherapy prior to tumour sampling. Records detailing systemic treatments received were obtained from the NHSD admitted patient care and outpatient tables, and the PHE-NCRAS AV Treatment and Systemic Anti-Cancer Therapy (SACT) tables. Records detailing radiotherapy received were obtained from the PHE-NCRAS AV Treatment and National Radiotherapy Dataset (RTDS) tables.

**Study sample selection**

Tumour samples were excluded if clinical data were missing or if unresolvable conflicts existed between clinical data sources (**Supplementary Table 1**). Overall, 2,251 of the 14,129 (15.9%) of tumour samples were excluded because: (1) Sex reported by PHE-NCRAS, NHSD and/or the GMC conflicted with the sex inferred from sequencing data; (2) It was not possible to assign the tumour to one of the 35 tumour groups, either because of missing or conflicting originating tissue or tumour histology data, or because the disease was not represented by one group; (3) It was unclear whether a primary tumour, a metastasis or a recurrence of a primary tumour was sampled due to missing or conflicting data; (4) It was not possible to determine the date of sampling due to missing or conflicting data; (5) Patient was <18 years old on the date of sampling.

Tumour sample purity and sequencing data quality affect variant calling precision and sensitivity and we therefore applied additional quality control criteria to the sequencing data (**Supplementary Table 1**)^1^. Overall, 267/11,878 (2.2%) of tumour samples with available clinical data were excluded based on the following sequencing data quality control criteria: (1) Tumour or matched germline sample cross-contamination was >1%, as determined by VerifyBamID^2^; (2) Number of SNVs called in a tumour was a low outlier for the associated tumour group - SNV number outliers were defined as tumors with a tumor-group-specific log-transformed SNV number Z-score < -3. To ensure that no individual was represented in the same tumour group, we removed duplicated samples from the same individual, preferentially keeping primary tumor samples of highest purity, as estimated by Ccube^3^. A further 505 non-solid tumour samples were excluded as these tumour types require complete copy number and structural variant calling to ascertain clinical actionability. 10,478 tumour samples were suitable for analysis following these criteria (**Supplementary Table 1**). The final cohort consisted of 9,693 primary tumours, 634 metastases and 151 primary tumour recurrences from 10,470 individuals. Eight patients were represented in more than one tumour group.

**Whole genome sequencing**

Paired tumour-normal tissue samples were obtained as part of the 100KGP cancer programme; an initiative for sequencing NHS patients diagnosed with cancer^4,5^. Recruitment of patients was through 13 Genomic Medicine Centres (GMCs) and affiliated hospitals. All patients provided written informed consent. Tissue collection and preparation, extraction and quantification of DNA was undertaken locally, and DNA transferred to a central national biorepository. Whole genome sequencing of paired tumour-germline DNA was conducted by Illumina. Additional processing, quality checking and data storage was performed by Genomics England.

Sample preparation was conducted using Illumina TruSeq DNA PCR-free library preparation kits. Sequencing was performed using HiSeq X, generating 150 base-pair paired-end reads. Tumour and germline samples were sequenced to average depths of 100x and 30x respectively. Poor sequencing quality outliers were identified using principal component analysis and excluded (based on the following quality metrics: average insert size, AT/CG dropout, unevenness of local coverage, percentage of mapped reads and percentage of chimeric DNA fragments). Sequencing quality outliers were not included in the 100KGP main programme releases and were therefore not considered in this study. Reads were aligned to the *Homo sapiens* GRCh38Decoy assembly using Isaac v03.16.02.19^6^. Paired tumour/germline sequencing data for 14,129 cancer samples were obtained from the 100KGP main program version 11 release.

**Somatic variant calling**

Somatic single nucleotide variant (SNV) and small insertion and deletion (indel) calling was performed using Strelka (v2.4.7)^7^. Variants were excluded if they failed any of the default Strelka filters or met any of the following criteria: (1) population germline allele frequency ≥1% in the gnomAD or 100KGP cohorts^8^; (2) somatic frequency ≥5% in 100KGP tumour samples; (3) overlapped a simple repeat as defined by Tandem Repeats Finder^9^; (4) indel is in a region with high levels of sequencing noise. High sequencing noise defined as ≥10% of base calls in a window extending 50 base pairs to either side of the indel call being excluded by Strelka.

SNV likely to be an artefact caused by systematic mapping or calling issues were identified by computing the ratio of tumour allele depths at each somatic SNV site and comparing against the ratio of allele depths at the same site in a panel of 7,000 normal germline samples. Allele depth at each site was counted, using bcftools mpileup function (version 1.9), considering only individuals not carrying the corresponding alternate allele. Duplicate reads were removed prior to counting and mapping quality ≥5 and base quality ≥5 thresholds were applied to replicate Strelka filters. SNVs with a Phred quality score ≤50 computed using Fisher’s exact test were excluded. Copy number alterations (CNAs) were called using Battenberg^10^ following variant allele frequency correction with alleleCount-FixVAF^11^.

**Annotation of mutations**

Somatic mutations were annotated to GRCh38 Ensembl v101 using the variant effect predictor (VEP)^12^. The following parameters were used: “vep -i <input_vcf> --assembly GRCh38 –no_stats –cache –offline –symbol –protein -o <output> --vcf –canonical –dir <ref_dir> --hgvs –hgvsg –fasta <GRCh38_fasta> --plugin CADD,<CADD_score_file> --plugin UTRannotator,<GRCh38_uORF_reference>”. The <CADD_score_file> was obtained using CADD v1.6 (<https://cadd.gs.washington.edu/>; with scores obtained for all SNV and indel mutations using the CADD software (https://github.com/kircherlab/CADD-scripts/), before being utilised by the VEP CADD plugin^13–15^. The plugin “UTRannotator” (https://github.com/ImperialCardioGenetics/UTRannotator) was used to annotate the potential impact of five prime untranslated region (5’ UTR) mutations^16^.

**Identification of driver genes**

Protein-coding driver genes were identified using the IntOGen pipeline (<https://bitbucket.org/intogen/intogen-plus/src/master/>; downloaded February 2021; https://intogen.readthedocs.io/en/latest/)^17^.

***Pre-processing of mutations -*** Somatic mutations passing filtering criteria described previously across a tumour cohort were subject to initial sample and mutation pre-processing. In the case of multiple tumours from the same patient, the primary tumour was used (in the case of primary/recurrence pair), alternatively the tumour with the highest purity was used. In each cohort, hypermutated tumours were flagged for exclusion from downstream driver gene identification if containing > 10,000 mutations and having an outlier mutation count (count > 1.5* interquartile range (IQR) + upper quartile (UQ)). Mutations found to be present in a Hartwig Panel of Normals were further excluded. Unless otherwise specified, mutations were mapped to canonical protein-coding transcripts from Ensembl v101.

***Running driver identification methods*** *-* Seven complementary driver gene identification methods were run: (1) dNdSCV^18^, for the Skin_Melanoma samples which contain a high proportion of hypermutated tumours, the parameter “max_coding_muts_per_sample = Inf” was used; (2) OncodriveFML^18,19^, CADD v1.6 scores were used as a measure of functional impact^15^^13^^14^; (3) OncodriveCLUSTL^20^, for the Skin_Melanoma samples, which contain a high proportion of hypermutated tumours, pentamer signatures were used rather than trinucleotide signatures; (4) cBaSE^21^; (5) MutPanning^22^; (6) HotMaps3D^23^; and (7) smRegions^24^, this analysis utilised information from protein family (pfam) domains, which were mapped to Ensembl v101 canonical transcripts.

***Combining driver identification methodologies*** *-* The driver combination procedure considered the top-100 ranked genes and their association *P* and *Q-*values in each of the seven driver identification methods. Briefly, genes assigned as Tier 1 or Tier 2 somatically mutated genes in the COSMIC Cancer Gene Census (<https://cancer.sanger.ac.uk/census>; v92 downloaded February 2021) were designated as “CGC” genes and represented a “truth” set of known drivers^25^. Through comparison of the relative enrichment of known drivers in the top ranked gene lists a per-method weighting was obtained. Per-method ranked lists were combined using Schulze’s voting method to generate a consensus ranking, with combined *P*-values estimated using a weighted Stouffer *Z*-score method.

Driver candidates were then classified into the following tiers: Tier 1 – Candidates where the consensus ranking is higher than the ranking of the first gene with stouffer *Q* > 0.05. These represent high confidence drivers; Tier 2 – Candidates not meeting the criteria for Tier 1 but which are CGC genes, and show a combined stouffer *Q_CGC_<*0.25. Representing a “rescue” of known cancer drivers; Tier 3 – Candidates not meeting the criteria for Tier 1 or Tier 2 but which have stouffer *Q* < 0.05. These represent lower confidence drivers; Tier 4 – Candidates not meeting criteria for Tier 1 or Tier 2 and stouffer *Q* > 0.05. These represent candidates that are not likely to be drivers.

***Post-processing of candidate drivers*** *-* Candidate driver genes were filtered on the basis of the following annotations:

1.	“AUTOMATIC FAIL” – a candidate driver gene would be excluded from further consideration if annotated by at least one of the following:

a. “TIER4” – categorised into Tier 4 by the combination procedure

b. “1_METHOD” – only significant (*Q*<0.1) in 1/7 methods (non-CGC genes)

c. “EXPRESSION” – gene has very low or absent expression in the relevant The Cancer Genome Atlas (TCGA) Tumor type

d. “OLFACTORY_RECEPTOR” – gene in list of olfactory receptor genes

e. “KNOWN_ARTIFACT” – gene is in a known list of artifacts or long genes (e.g. TTN)

2.	“MANUAL REVIEW” – if a gene is not excluded on the basis of any “AUTOMATIC FAIL” filters, it is retained as a candidate driver

a. “GERMLINE” – non-Tier 1-CGC gene has 1+ mutations per sample and oe_syn/ms/lof > 1.5 based on GnomAD v2.1 constraint metric estimates (https://gnomad.broadinstitute.org/downloads#v2-constraint)

b. “SAMPLE_3_MUTS” – non-CGC gene where there are 3+ mutations in 1+ Tumor

c.	“LITERATURE” – non-CGC gene where there are no literature annotations according to CancerMine (http://bionlp.bcgsc.ca/cancermine/; downloaded February 2021)^26^.

3.	“AUTOMATIC PASS” – is not flagged by any “AUTOMATIC FAIL” or “MANUAL REVIEW” filters

Candidate driver roles were assigned on the basis of the dN/dS ratios for missense (*wmis*) and nonsense (*wnon*) mutations for the given gene derived from dNdSCV (<https://bitbucket.org/intogen/intogen-plus/src/master/core/intogen_core/postprocess/drivers/role.py>): a “distance” metric was calculated by $\frac{(wmis-wnon)}{\sqrt{2}};$candidate drivers where $distance>0.1$ represent those with an excess of missense to nonsense mutations and are assigned as “Oncogenes”; candidate drivers where $distance < 0.1$ represent those with an excess of nonsense to missense mutations and are assigned as “Tumor suppressor genes (TSGs)”; otherwise, the role of the candidate driver is unclear and was assigned as “Ambiguous”.

In the case of multiple cohorts being run representing subsets of a given tumour type, a “consensus” role was designated comparing between each subtype role (“Oncogene” if 1+ cohort and no other cohorts assigned as “TSG”, “TSG” if in 1+ cohort and no other cohorts assigned as “Oncogene”, otherwise “ambiguous”).

Gene candidates were annotated by their overlap with any IntOGen cohorts from a previous 2020.02.01 pan-cancer analysis (<https://www.intogen.org/download?file=IntOGen-Cohorts-20200201.zip>) as well as from a pan-cancer TCGA analysis Bailey et al., 2018^27^.

**Mutations exhibiting extreme strand bias**

SNV mutations that otherwise pass filtering criteria as previously detailed were scrutinised if they showed excessive strand bias (Strelka INFO field “SNVSB=” >10). This highlighted the large number of mutations causing the missense change in *CACNA1E (p.Ile95Leu*), which showed excessive strand bias and is therefore likely to be false-positive. Hence, it is likely that *CACNA1E* itself is likely to be a false-positive driver candidate. In addition we identified WDR64, VCP, GOLGA6L10, NBPF1, TUBB8, TRIM64B, HLA-DQB2 and KIR3DL2 genes as likely false-positives on this basis (**Supplementary Table 14**).

**OncoKB annotation of driver mutations**

Nonsynonymous mutations in the 684 gene transcripts considered by OncoKB v3.11 were annotated using the OncoKB API (<https://www.oncokb.org/>)^28^. In the first instance, the HGVSg identifier was used, however in rare instances if this failed a combination of gene symbol, consequence and HGVSp were used to map mutations to OncoKB annotations.

***Annotation of oncogenic mutations -*** Nonsynonymous mutations in candidate driver genes were annotated as “Oncogenic” if either of the following criteria were met: (1) the mutation is annotated by OncoKB as “Oncogenic”, “Likely Oncogenic” or “Predicted Oncogenic”; (2) the driver role is “Oncogene”, consequence is “missense” and mutation is recurrent (seen in >0.5% Tumors pan-cancer); (3) the driver role is “TSG” or “ambiguous” and either the consequence is protein-truncating (“splice acceptor”, “splice donor”, “frameshift”, “stop lost”, “stop gained”, “start lost”) or “missense” and mutation is recurrent (seen in >0.5% Tumors pan-cancer). For POLE, oncogenic annotations were restricted to missense mutations in the exonuclease domain (amino acid residues 268-471). Nonsynonymous mutations not meeting these criteria were considered as a variant of uncertain significance (VUS).

***Lollipop plots of driver gene mutations -*** Lollipop plots of driver gene mutations were generated using the R package trackViewer (<https://github.com/jianhong/trackViewer>^29^. Pfam protein domains mapping to the Ensembl v101 canonical transcripts were plotted. The protein position was taken from the first position in the HGVSp annotation, other than for splice donor and acceptor mutations where the codon nearest to the HGVSc transcript position was assigned as the protein position.

***Power estimates for driver gene estimation -*** Estimates of power to detect a driver assumes a driver gene has a higher non-silent mutation rate compared to the corresponding background mutation rate^30^. A binomial model is used to theoretically compare a driver gene with a non-driver conditional on the sample size. The mutation rate factor, $F_{g}$= 3.9, is defined such that driver genes in the 90th percentile of gene-specific background mutation rates (as calculated by MutsigCV^31^) are included. Letting $\theta$ be the exome-wide background mutation rate (mutations per base) of a tumour cohort, the general gene mutation background rate for the 90th percentile of genes is $\mu=F_{g}\theta$. $r$ is the non-silent mutation rate above the background rate. The effective gene length is defined as $L_{EFF}=3L/4$ assuming the ratio of non-silent to silent mutations is $3$ to $1$ and $L=1,500$ represents the average gene length. The power from the binomial model considers the hypotheses; $H_{0}: \mu_{NULL}=F_{g}\theta$, vs $H_{1}: \mu_{ALT}=1-((1-\mu_{NULL})^{L_{EFF}} -r)^{(1/L_{EFF})}$, where $\mu_{ALT}$ is the non-silent mutation rate of a suspected driver gene. The null hypothesis is rejected at a *P*-value of $5\times{10}^{-6}$. If there is enough power for 90% of genes, thus we consider the mutation rate for the 90th percentile of genes to be classified as a driver via binomial probability, then the IntOGen pipeline is likely to have sufficient power too.

**Comparison of driver rates in histologies**

To compare the rates of driver somatic mutations in different histologies per cancer tissue, expected mutation rate and uncertainties were estimated by taking a uniform distribution prior with a binomial likelihood function with the number of samples which are mutated ($k$) vs total samples ($n$) for the given cancer site. This produces a beta distribution posterior with parameters $\alpha=k+1$, $\beta=n-k+1$ such that the expected mutation rate is $(k+1)/(n+2)$. For each histology, *P*-values are evaluated using a binomial test of the mutation rate of the samples with the given histology against the mutation rate of all samples for the given organ.

**Assessment of domain specific mutations**

We assessed domain specific mutations by considering the cancer drivers where smRegions is a significant bidder (*Q*-value <0.1) and the driver is annotated in multiple cancer types.

**Predicting mismatch repair deficiency**

Samples with evidence of defective mismatch repair were detected using MSINGS, following the previously described procedure for background model generation^32,33^.

**Copy number alteration profiling**

Clonal and sub-clonal somatic copy number alterations (CNAs) were detected using an iterative procedure incorporating Battenberg v2.2.8 (**Supplementary Figure 16**)^10^. This procedure comprises four stages: (stage 1) initial CNA profiling; (stage 2) evaluation of CNA profile concordance with variant allele frequency (VAF) distributions; (stage 3) quality assessment; and for those samples that fail quality assessment, (stage 4) CNA re-profiling with alternative purity and ploidy estimates and repeat of stages 2 and 3.

*Stage 1: Initial profiling of copy number alterations*

Battenberg was used to detect clonal and sub-clonal CNAs and to estimate sample purity and tumour ploidy^10^. Briefly, numbers of reads supporting single-nucleotide polymorphism (SNP) reference and alternate alleles were counted in tumour and normal samples using alleleCount-FixVAF, which removes reference bias introduced by ISAAC^11^. Heterozygous SNPs were phased using SHAPEIT2 v2.r904 and A and B alleles assigned^34^. These data were then segmented using piece-wise constant fitting and sub-clonal copy-number segments identified using t-tests^35^. Sample purity and tumour ploidy were estimated using the method described by Van Loo et al. (2010)^36^. As sequencing data were aligned to hg38, SNP positions were converted to hg37 before phasing, and output segments converted back to hg38.

*Stage 2: Evaluation of profile concordance with variant allele frequency distributions*

Expected variant allele frequency is dependent on the fraction of tumour cells containing the variant, the tumour copy number profile, the number of chromosome copies with the variant (its multiplicity) and the sample purity^37^. Given the tumour copy number profile and an estimated sample purity, we can therefore expect to observe enrichment of variants with allele frequencies approximating particular values (representing variants present in all tumour cells)^36^. Failure to observe such enrichment would suggest that either the copy number profile or sample purity is incorrect, and we therefore assessed Battenberg output validity using SNV VAF distributions (**Supplementary Figure 17**).

When evaluating SNV VAF distributions, only autosomal genome segments with copy number states of 1:1, 1:0, 2:2, 2:1, 2:0 and no evidence of sub-clonal copy number changes were considered. Each of these five copy number states was evaluated separately, as the possible variant multiplicities and expected clonal SNV VAFs differ between them (**Supplementary Figure 17a**). Copy number states corresponding to genome regions containing <5% of all SNVs were not considered. Expected locations of VAF distribution peaks were computed as:

$\frac{\rho_{Battenberg}m}{2(1-\rho_{Battenberg}) + \rho_{Battenberg}\psi_{v}}$

Where r_Battenberg_ is the sample purity estimated by Battenberg, y*_v_* the ploidy of the tumour at the variant site, and *m* the variant multiplicity, which can equal 1 or 2 in copy number states of 2:2, 2:1 and 2:0 and only 1 in states of 1:1 and 1:0 (**Supplementary** **Figure 17b**). VAF distribution peaks were called using kernel density estimation, implemented in peakPick v0.11, with peaks corresponding to densities <0.3 being excluded (**Supplementary** **Figure 17c**)^38^. For each copy number state, the expected peak location corresponding to the highest considered variant multiplicity was matched to the observed VAF distribution peak with the greatest VAF, whilst any other expected peak location was matched to the observed peak with the most similar VAF (**Supplementary** **Figure 17d**). Tumour heterogeneity can prevent VAF peak detection, and therefore for samples where ≥1 expected peak locations were considered, the expected peak furthest from the respective matched observed peak (in terms of VAF) was discarded (**Supplementary** **Figure 17e**). Sample purity (r_i_) was re-estimated for each remaining expected peak location using the matched observed peak VAF (**Supplementary** **Figure 17f**):

${\rho_{i}=\frac{2\alpha}{m +\omega(2-\psi_{s})}}$

Where w is the VAF of the matched observed peak and y*_s_* the ploidy of the respective copy number state. Greater variant numbers improve our ability to call peaks, and therefore a single new purity estimate (r_new_) was computed as the weighted average of the peak-wise purity estimates (and used when re-profiling samples) (**Supplementary** **Figure 17g**):

${\rho_{new}=\sum_{i} \frac{n_{i}\rho_{i}}{Nq_{i}}}$

Where n_i_ is the number of SNVs in genome regions of the respective copy number state, N is the number of SNVs in genome regions of all considered copy number states, and q_i_ is the number of considered variant multiplicities for the respective copy number state. When assessing CNA profile quality, the weighted average of the difference between the purity estimated by Battenberg and the peak-wise purity estimates was used:

${n=\sum_{i} \frac{n_{i}{|\rho}_{i} - \rho_{Battenberg}|}{Nq_{i}}}$

*Stage 3: Quality assessment*

Multiple criteria were used to assess Battenberg output validity:

·	VAF distribution peaks observed at the correct locations (defined as h<5%).

·	DPClust identified a clonal mutation cluster (defined as a cluster containing ≥5% of all SNVs with a CCF of between 0.9 and 1.1)^10^.

·	DPClust identified no “super-clonal” mutation clusters (defined as clusters containing ≥5% of all SNVs with CCFs >1.1).

·	Where Battenberg identifies most of the genome to be tetraploid (2:2), a peak in the SNV VAF distribution in 2:2 regions corresponding to a variant multiplicity of 1 is observed.

·	No single homozygous deletion >10Mb is called.

Samples satisfying all criteria were deemed to pass and their CNA profiles and purity estimates were used in subsequent analyses. Samples not satisfying at least one criterion were deemed to fail and were re-profiled (*i.e.* proceeded to stage 4).

*Stage 4: Re-profiling of copy number alterations*

Samples failing quality assessment were re-profiled a maximum of three times using Battenberg with new purity and ploidy estimates. Samples failing quality assessment after three re-profiling attempts were not considered in subsequent analyses. The new purity (r_new_) was estimated in stage 2, whilst the new ploidy (y_new_) was estimated using the method described by Van Loo *et al.* (2010)^36^:

**Structural variants calling**

We identified structural variants (SVs; also referred to as rearrangements) using a graph-based consensus approach including Delly, Lumpy and Manta, and support from CNAs^39–41^. Delly was run with post-filtering of somatic SVs using all normal samples, and Lumpy and Manta were run with default parameters. Rearrangements from the three SV callers were excluded if <2% of tumour reads at the rearrangement breakpoint site supported the rearrangement, if any reads in the matched normal supported the rearrangement, or if either rearrangement breakpoint was in a centromeric or telomeric region, or on a non-standard reference contig (not chromosomes 1-22, X or Y). Remaining rearrangements were merged using a modified version of PCAWG Merge SV, which is a graph-based approach that identifies and merges rearrangements from multiple callers, allowing 400bp slop for ambiguity in rearrangement breakpoint position^42^. Rearrangements were included in the final call set if they were identified by at least two SV callers, or by one SV caller but with a rearrangement breakpoint <3kb from a CNA segment boundary.

xTea was used to identify somatically acquired long interspersed nuclear element (LINE-1) retrotransposition events^43^. Other retrotransposition categories, including Alu elements, SINE-VNTR-Alu elements and processed pseudogene, were not considered as they collectively comprise ≤3% of retrotransposition events across human cancers^44^. Retrotransposition and other SV-generating events are mechanistically distinct and we therefore excluded retrotransposition events from our SV analyses. SVs identified using the graph-based consensus approach were classified as potential retrotransposition events and excluded if: (1) xTea identified a transduced region within 10kb of either rearrangement break point in ≥1% of glioma samples, or (2) xTea identified a transduced region within 10kb of either rearrangement break point in the same sample. A threshold of 10kb was chosen as most somatically acquired transductions span regions <10kb from a canonical LINE-1 element^45^.

**Predicting homologous recombination deficiency**

Evidence of homologous recombination deficiency (HRD) was assessed using HRDetect^46^. HRDetect requires CNA data and was therefore run only on 9,207 tumours passing CNA calling. To compute the HRDetect score we determined the following input features: exposures of single base substitution signatures, SBS3 and SBS8, as well as COSMIC rearrangement signatures 3 and 5, the proportion of short deletions at microhomology, and the HRD-LOH index^46,47^. SBS3 and SBS8 contribution estimates were obtained from SigProfiler^48^. We used a probabilistic cutoff of 0.7, which translates to 98.7% sensitivity for predicting BRCA1/BRCA2 deficiency and has a high efficacy when applied to multiple cancer types^46^.

**Comparison of driver mutation frequencies between Genomics England and MSK**

To determine the sensitivity of WGS data from the 100kGP we first compared driver mutation frequencies with those from MSK-IMPACT and MSK-MET, a combined cohort of ~25,000 cancer patients whose tumours have been panel sequenced to identify driver mutations^49,50^. Mutation and sample data were downloaded from https://cbioportal-datahub.s3.amazonaws.com/msk_impact_2017.tar.gz and https://cbioportal-datahub.s3.amazonaws.com/msk_met_2021.tar.gz. Where possible, MSK tumours were matched to 100kGP tumour groupings on the basis of their oncotree code. For CNS tumours, IDH mutation status was used to classify GBM tumours as GBM, IDHwt or GBM, IDmut. Due to lack of 1p/19q co-deletion status, CNS tumours classified as mixed oligoastrocytomas were excluded. To maintain consistency with 100kGP, MSK mutations were lifted over to GRCh38 and annotated by VEP v101 and OncoKB. Samples were aggregated by tumour group and type (metastasis/primary) and the fraction of samples with an oncogenic mutation in a given driver gene were compared.

**Assessing the sensitivity of WGS to detect driver mutations**

By analysing the distribution of allelic depths in called PASS mutations in the 100kGP we found that the rate of calls falls when fewer than 6 reads support the alternate allele (**Supplementary Figure 6**). We therefore used 6 reads as a minimum coverage threshold to approximate the sensitivity of the WGS samples to somatic mutations. Per-base coverage was extracted from tumour bam files using GATK v4.4.0.0 DepthOfCoverage^51^. A gene panel of 43 representative driver genes was obtained from NHS Genomic Test Directory for Cancer (2021-22 v5.0 published 31 October 2022). Genomic regions were defined as per driver gene identification (*i.e.* coding sequence (CDS) from the canonical ensembl v101 transcript including essential splice sites). The TERT promoter region was defined as GRCh38 chr5:1295019-1295268. Coverage was mapped to gene panel regions using bedops v2.4.26 and bedtools v2.3.0 to obtain per-gene coverage statistics^52,53^. Assuming a heterozygous clonal mutation the expected number of reads is given by $0.5 \times coverage \times purity$. The distribution of coverage across samples for three common pancancer drivers is given in **Supplementary Figure 7**. The sensitivity is the probability of measuring at least 6 reads given the expected number of reads which we assume is Poisson distributed. We estimated this for all genes and samples shown in **Supplementary Figures 8 & 9**. In a realistic worst case, for a read coverage of 75 (which is in the lower 5th percentile of genes in 100kGP samples) with a tumour purity of 0.2, the sensitivity is 76% however this rises to 99.98% for a purity of 0.5. We also estimated the fraction of each driver gene with an expected alt read count greater than 6. Results for TP53, KRAS and PIK3CA are given in **Supplementary Figure 10**.

**Comparison of actionability between WGS and panel**

Actionable driver genes were defined from the OncoKB and COSMIC databases. This list compared with the NHS Genomic Test Directory for Cancer (2021-22 v5.0 published 31 October 2022) at a mutation-specific level.

**Timing driver mutations**

The relative evolutionary timing of candidate driver mutations was obtained using MutationTimeR (<https://github.com/gerstung-lab/MutationTimeR>)^54^.

*Preparing MutationTimeR input files*

Copy number input for MutationTimeR was prepared from Battenberg segmentation files, with the clonal frequency of each segment taken as the tumour purity. In the case of subclonal calls, the clonal frequency was calculated by multiplying the tumour purity by the clonal fraction.

The clusters input for MutationTimeR was prepared from DPClust cluster estimates. The VAF proportion was calculated by multiplying the estimated cluster CCF by the tumour purity. Superclonal clusters (CCF>1.1) were removed.

VCF input for MutationTimeR was obtained from the small somatic SNV/indel variant VCFs which had been filtered as previously described. For SNVs, alt and ref depths were obtained using FixVAF (<https://github.com/danchubb/FixVAF>). For indels, ref and alt depths were obtained from Tier2 Strelka TAR and TIR fields respectively. Only mutations within Battenberg copy-number segments were retained (note: for male XY tumours with only 1 copy of the X chromosome copy number information is restricted to the pseudoautosomal region (PAR) and battenberg was not run on the Y chromosome).

*Running MutationTimeR*

MutationTimeR was run with 1,000 bootstraps. For tumours previously defined as having undergone whole genome doubling (WGD), the parameter “isWgd” was set to TRUE. Mutations were then classified into estimated simple clonal states (as per Fig.1a of Gerstung *et al.* (2020)^54^):

- “Clonal [early]” – Mutation on ≥ 2 copies per cell
- “Clonal [late]” – Mutation on 1 copy per cell, no retained allele
- “Clonal [NA]” – Mutation on 1 copy per cell, either on amplified or retained allele
- “Subclonal” – Mutation on < 1 copy per cell

**REFERENCES**

1.	Saunders, C. T. *et al.* Strelka: accurate somatic small-variant calling from sequenced tumor-normal sample pairs. *Bioinformatics* **28**, 1811–1817 (2012).

2.	Jun, G. *et al.* Detecting and estimating contamination of human DNA samples in sequencing and array-based genotype data. *Am. J. Hum. Genet.* **91**, 839–848 (2012).

3.	Yuan, K., Macintyre, G., Liu, W., Markowetz, F. & PCAWG-11 working group. Ccube: A fast and robust method for estimating cancer cell fractions. Preprint at https://doi.org/[10.1101/484402](http://dx.doi.org/10.1101/484402).

4.	Turnbull, C. Introducing whole-genome sequencing into routine cancer care: the Genomics England 100 000 Genomes Project. *Ann. Oncol.* **29**, 784–787 (2018).

5.	Turnbull, C. *et al.* The 100 000 Genomes Project: bringing whole genome sequencing to the NHS. *BMJ* k1687 Preprint at https://doi.org/[10.1136/bmj.k1687](http://dx.doi.org/10.1136/bmj.k1687) (2018).

6.	Raczy, C. *et al.* Isaac: ultra-fast whole-genome secondary analysis on Illumina sequencing platforms. *Bioinformatics* **29**, 2041–2043 (2013).

7.	Kim, S. *et al.* Strelka2: fast and accurate calling of germline and somatic variants. *Nat. Methods* **15**, 591–594 (2018).

8.	Karczewski, K. J. *et al.* The mutational constraint spectrum quantified from variation in 141,456 humans. *Nature* **581**, 434–443 (2020).

9.	Benson, G. Tandem repeats finder: a program to analyze DNA sequences. *Nucleic Acids Research* vol. 27 573–580 Preprint at https://doi.org/[10.1093/nar/27.2.573](http://dx.doi.org/10.1093/nar/27.2.573) (1999).

10.	Nik-Zainal, S. *et al.* Mutational processes molding the genomes of 21 breast cancers. *Cell* **149**, 979–993 (2012).

11.	Cornish, A. J. *et al.* Reference bias in the Illumina Isaac aligner. *Bioinformatics* vol. 36 4671–4672 (2020).

12.	McLaren, W. *et al.* The Ensembl Variant Effect Predictor. *Genome Biol.* **17**, 122 (2016).

13.	Rentzsch, P., Witten, D., Cooper, G. M., Shendure, J. & Kircher, M. CADD: predicting the deleteriousness of variants throughout the human genome. *Nucleic Acids Res.* **47**, D886–D894 (2019).

14.	Rentzsch, P., Schubach, M., Shendure, J. & Kircher, M. CADD-Splice-improving genome-wide variant effect prediction using deep learning-derived splice scores. *Genome Med.* **13**, 31 (2021).

15.	Kircher, M. *et al.* A general framework for estimating the relative pathogenicity of human genetic variants. *Nat. Genet.* **46**, 310–315 (2014).

16.	Zhang, X., Wakeling, M., Ware, J. & Whiffin, N. Annotating high-impact 5’untranslated region variants with the UTRannotator. *Bioinformatics* **37**, 1171–1173 (2021).

17.	Martínez-Jiménez, F. *et al.* A compendium of mutational cancer driver genes. *Nat. Rev. Cancer* **20**, 555–572 (2020).

18.	Martincorena, I. *et al.* Universal Patterns of Selection in Cancer and Somatic Tissues. *Cell* **171**, 1029–1041.e21 (2017).

19.	Mularoni, L., Sabarinathan, R., Deu-Pons, J., Gonzalez-Perez, A. & López-Bigas, N. OncodriveFML: a general framework to identify coding and non-coding regions with cancer driver mutations. *Genome Biol.* **17**, 128 (2016).

20.	Arnedo-Pac, C., Mularoni, L., Muiños, F., Gonzalez-Perez, A. & Lopez-Bigas, N. OncodriveCLUSTL: a sequence-based clustering method to identify cancer drivers. *Bioinformatics* **35**, 4788–4790 (2019).

21.	Weghorn, D. & Sunyaev, S. Bayesian inference of negative and positive selection in human cancers. *Nat. Genet.* **49**, 1785–1788 (2017).

22.	Dietlein, F. *et al.* Identification of cancer driver genes based on nucleotide context. *Nat. Genet.* **52**, 208–218 (2020).

23.	Tokheim, C. *et al.* Exome-Scale Discovery of Hotspot Mutation Regions in Human Cancer Using 3D Protein Structure. *Cancer Res.* **76**, 3719–3731 (2016).

24.	Porta-Pardo, E. & Godzik, A. e-Driver: a novel method to identify protein regions driving cancer. *Bioinformatics* **30**, 3109–3114 (2014).

25.	Sondka, Z. *et al.* The COSMIC Cancer Gene Census: describing genetic dysfunction across all human cancers. *Nat. Rev. Cancer* **18**, 696–705 (2018).

26.	Lever, J., Zhao, E. Y., Grewal, J., Jones, M. R. & Jones, S. J. M. CancerMine: a literature-mined resource for drivers, oncogenes and tumor suppressors in cancer. *Nat. Methods* **16**, 505–507 (2019).

27.	Bailey, M. H. *et al.* Comprehensive Characterization of Cancer Driver Genes and Mutations. *Cell* **173**, 371–385.e18 (2018).

28.	Chakravarty, D. *et al.* OncoKB: A Precision Oncology Knowledge Base. *JCO Precis Oncol* **2017**, (2017).

29.	Ou, J. & Zhu, L. J. trackViewer: a Bioconductor package for interactive and integrative visualization of multi-omics data. *Nat. Methods* **16**, 453–454 (2019).

30.	Tokheim, C. J., Papadopoulos, N., Kinzler, K. W., Vogelstein, B. & Karchin, R. Evaluating the evaluation of cancer driver genes. *Proc. Natl. Acad. Sci. U. S. A.* **113**, 14330–14335 (2016).

31.	Lawrence, M. S. *et al.* Discovery and saturation analysis of cancer genes across 21 tumour types. *Nature* **505**, 495–501 (2014).

32.	Cornish, A. J. *et al.* Whole genome sequencing of 2,023 colorectal cancers reveals mutational landscapes, new driver genes and immune interactions. *bioRxiv* 2022.11.16.515599 (2022) doi:[10.1101/2022.11.16.515599](http://dx.doi.org/10.1101/2022.11.16.515599).

33.	Salipante, S. J., Scroggins, S. M., Hampel, H. L., Turner, E. H. & Pritchard, C. C. Microsatellite instability detection by next generation sequencing. *Clin. Chem.* **60**, 1192–1199 (2014).

34.	Delaneau, O., Marchini, J. & Zagury, J.-F. A linear complexity phasing method for thousands of genomes. *Nat. Methods* **9**, 179–181 (2011).

35.	Nilsen, G. *et al.* Copynumber: Efficient algorithms for single- and multi-track copy number segmentation. *BMC Genomics* **13**, 591 (2012).

36.	Van Loo, P. *et al.* Allele-specific copy number analysis of tumors. *Proc. Natl. Acad. Sci. U. S. A.* **107**, 16910–16915 (2010).

37.	Dentro, S. C., Wedge, D. C. & Van Loo, P. Principles of Reconstructing the Subclonal Architecture of Cancers. *Cold Spring Harb. Perspect. Med.* **7**, (2017).

38.	Weber, C. M., Ramachandran, S. & Henikoff, S. Nucleosomes are context-specific, H2A.Z-modulated barriers to RNA polymerase. *Mol. Cell* **53**, 819–830 (2014).

39.	Rausch, T. *et al.* DELLY: structural variant discovery by integrated paired-end and split-read analysis. *Bioinformatics* **28**, i333–i339 (2012).

40.	Layer, R. M., Chiang, C., Quinlan, A. R. & Hall, I. M. LUMPY: a probabilistic framework for structural variant discovery. *Genome Biol.* **15**, R84 (2014).

41.	Chen, X. *et al.* Manta: rapid detection of structural variants and indels for germline and cancer sequencing applications. *Bioinformatics* **32**, 1220–1222 (2016).

42.	Li, Y. *et al.* Author Correction: Patterns of somatic structural variation in human cancer genomes. *Nature* **614**, E38 (2023).

43.	Chu, C. *et al.* Comprehensive identification of transposable element insertions using multiple sequencing technologies. *Nat. Commun.* **12**, 3836 (2021).

44.	Rodriguez-Martin, B. *et al.* Pan-cancer analysis of whole genomes identifies driver rearrangements promoted by LINE-1 retrotransposition. *Nat. Genet.* **52**, 306–319 (2020).

45.	Tubio, J. M. C. *et al.* Mobile DNA in cancer. Extensive transduction of nonrepetitive DNA mediated by L1 retrotransposition in cancer genomes. *Science* **345**, 1251343 (2014).

46.	Davies, H. *et al.* HRDetect is a predictor of BRCA1 and BRCA2 deficiency based on mutational signatures. *Nat. Med.* **23**, 517–525 (2017).

47.	Nik-Zainal, S. *et al.* Landscape of somatic mutations in 560 breast cancer whole-genome sequences. *Nature* **534**, 47–54 (2016).

48.	Ashiqul Islam, S. M. *et al.* *Uncovering Novel Mutational Signatures by de Novo Extraction with SigProfilerExtractor*. (2022).

49.	Zehir, A. *et al.* Mutational landscape of metastatic cancer revealed from prospective clinical sequencing of 10,000 patients. *Nat. Med.* **23**, 703–713 (2017).

50.	Nguyen, B. *et al.* Genomic characterization of metastatic patterns from prospective clinical sequencing of 25,000 patients. *Cell* **185**, 563–575.e11 (2022).

51.	McKenna, A. *et al.* The Genome Analysis Toolkit: a MapReduce framework for analyzing next-generation DNA sequencing data. *Genome Res.* **20**, 1297–1303 (2010).

52.	Neph, S. *et al.* BEDOPS: high-performance genomic feature operations. *Bioinformatics* **28**, 1919–1920 (2012).

53.	Quinlan, A. R. & Hall, I. M. BEDTools: a flexible suite of utilities for comparing genomic features. *Bioinformatics* **26**, 841–842 (2010).

54.	Gerstung, M. *et al.* The evolutionary history of 2,658 cancers. *Nature* **578**, 122–128 (2020).
